# Distinct pathway-based effects of blood pressure and body mass index on cardiovascular traits: comparison of novel Mendelian Randomization approaches

**DOI:** 10.1101/2023.10.31.23297593

**Authors:** Maria K Sobczyk, Tom G Richardson, Genevieve M Leyden, Tom R Gaunt

## Abstract

**Background:** Mendelian randomization (MR) leverages genetic variants as instrumental variables to determine causal relationships in epidemiology. However, challenges persist due to heterogeneity arising from horizontal pleiotropy. On the other hand, exploration of the biological underpinnings of such heterogeneity across variants can enhance our understanding of disease mechanisms and inform therapeutic strategies. Here, we introduce a new approach to instrument partitioning based on enrichment of Mendelian disease categories and compare it to a method based on genetic colocalisation in contrasting tissues.

**Methods:** We employed one-sample and two-sample MR methodologies using blood pressure (BP) exposure SNPs grouped by proximity to Mendelian disease genes affecting the renal system or vasculature, or body mass index (BMI) variants related to mental health and metabolic Mendelian disorders. We then compared the causal effects of Mendelian-partitioned SNPs on cardiometabolic outcomes with subsets inferred from gene expression colocalisation in kidney, artery (for BP), adipose, and brain tissues (for BMI). Additionally, we assessed whether effects from these groupings could emerge by chance using random SNP subset sampling.

**Results:** Our findings suggest that the causal relationship between systolic BP and coronary heart disease is predominantly driven by SNPs associated with vessel-related Mendelian diseases over renal. However, kidney-oriented SNPs showed more pronounced effect size in the colocalization-based analysis, hinting at a multifaceted interplay between pathways in the disease aetiology. We consistently identified a dominant role of Mendelian vessel and coloc artery exposures in driving the negative effect of diastolic BP on left ventricular stroke volume and positive effect of systolic BP on type 2 diabetes. We also found higher causal estimates for metabolic versus mental health SNPs when dissecting BMI pathway contribution to atrial fibrillation risk using Mendelian disease. In contrast, brain variants yielded higher causal estimates than adipose in the colocalization method.

**Conclusions:** This study presents a novel approach to dissecting heterogeneity in MR by integrating clinical phenotypes associated with Mendelian disease. Our findings emphasize the importance of understanding tissue-/pathway- specific contributions in interpreting causal relationships in MR. Importantly, we advocate caution in interpreting pathway-partitioned effect size differences without robust statistical validation.

## Introduction

Mendelian randomization (MR) is a statistical method used in epidemiology to study the causal relationship between a risk factor (exposure) and an outcome (disease or trait) by leveraging genetic variants derived from genome-wide association studies (GWAS) as instrumental variables^1^. The technique is based on the principles of Mendelian inheritance, which states that genetic variants, such as single nucleotide polymorphisms (SNPs), are randomly assigned during meiosis and therefore should be less prone to confounding factors or reverse causation that typically plague observational studies (subject to meeting certain assumptions^2^).

However, causal estimates derived from individual genetic variants used as instrumental variables in MR studies can be highly variable^3^. Heterogeneity in a given single result can arise from two main sources of bias due to violation of key MR assumptions. These are horizontal pleiotropy, which occurs when one or more genetic variants influence the outcome through multiple independent pathways and weak instrument bias which can result in imprecise causal estimates and increase heterogeneity^4^.

Of chief interest in our study, causal effects vary due to endpoint phenotypes being de facto composites, representing divergent underlying biological mechanisms covered by different genetic instruments, which unlike bias can improve understanding of disease aetiology and help design better targeted interventions. Three broad types of approaches have been used so far when studying biological sources of heterogeneity: direct clustering based on SNP associations with exposure and outcome^5,6^, clustering of variant associations across a set of traits^7–9^, or instrument clustering informed by tissue gene expression patterns^10–13^. In particular, a biological hypothesis-driven approach proposed by Leyden et al. (2022)^11^ clusters genetic instruments for body mass index (BMI) based on the tissue (brain or adipose) where a given BMI SNP is found to colocalise with an expression quantitative trait locus (eQTL). The Bayesian colocalisation method coloc^14^ is employed here to compare the association signals at a specific genomic region for the two traits of interest (gene eQTL and BMI), considering variables such as effect sizes and allele frequencies to determine to what extent these are consistent with a single shared causal variant for both traits. In this way, a given SNP instrument is putatively linked to a particular gene whose expression (either in the adipose or brain tissue, or both) potentially mediates distinct causal effect on a set of cardiometabolic traits.

Although this approach was used to prioritise the putative causal tissue types underlying BMI-associated genes, in general the coloc method as originally implemented has been shown to lack specificity when assigning SNPs to genes on its own, particularly when using eQTL data due to the co-expression of nearby genes^15^. Another approach of prioritizing candidate genes at GWAS loci is to leverage the knowledge of Mendelian monogenic diseases, which are caused by rare mutations with large effects on phenotypes. Several studies have reported an enrichment of Mendelian disease genes near GWAS loci across various phenotypes, suggesting shared genetic basis between complex and Mendelian traits^16–18^.

However, not all Mendelian disease genes are equally relevant for a given complex trait, and trait symptom similarity ought to be a key metric for gene prioritization^19^.

In this paper, we introduce a new approach of Mendelian disease category-driven stratification of variants for common exposures used in MR. Blood pressure (BP) is a highly polygenic risk factor for a number of cardiovascular^20–22^ and metabolic^23–25^ conditions, with both vasculature-^26–28^ and kidney-^29–31^ expressed genes shown previously to be of key importance. Kidneys control blood pressure by regulating blood volume and electrolyte balance^32^, chiefly through natriuresis response^33^ and the renin-angiotensin-aldosterone system (RAAS) hormonal axis. Accordingly, impaired kidney function has long been linked to hypertension^34^. The vasculature regulates blood pressure via modulation of vascular tone. This is achieved through the processes of vasoconstriction and vasodilation, controlled by the smooth muscle cells in the arterial walls^35^. The endothelium lining the inner surface of blood vessels plays a pivotal role by releasing an array of vasoactive substances^36^. Endothelial cells secrete endothelin, a potent vasoconstrictor and nitric oxide, the key vasodilator^37,38^ and can also influence blood pressure through inflammatory mechanisms^39^. If instruments acting on BP via these two key mechanisms show different estimates of effect on an outcome in MR, we hypothesize that this will be due to pleiotropy in one or both subsets of instruments.

We begin by contrasting the kidney and vascular components of blood pressure risk factor burden on cardiometabolic disease. To achieve this, we carry out one-sample and two-sample multivariable MR analyses utilising blood pressure (systolic and diastolic) exposure variants grouped by co-sharing genetic loci with Mendelian disease genes whose symptoms affect either the renal system or vasculature (**Figure 1**). We compare our results to the colocalisation-based method proposed by Leyden et al. (2022)^11^ by linking blood pressure genetic variants to regulation of gene expression in kidney or arteries. We then return to the BMI exposure reported by Leyden et al.^11^ to ask if the effect of variants grouped by link to metabolic and mental health Mendelian disease corresponds to the effect obtained by adipose and brain tissue colocalisation-based subsets. Finally, using random re-sampling of the exposure SNP sets we investigate if the effect size differences observed in coloc- or Mendelian- based SNP subdivisions are likely to arise by chance. Our findings provide valuable insights into tissue-based underpinnings of causal links between BP or BMI and cardiometabolic traits.

**Figure 1.**
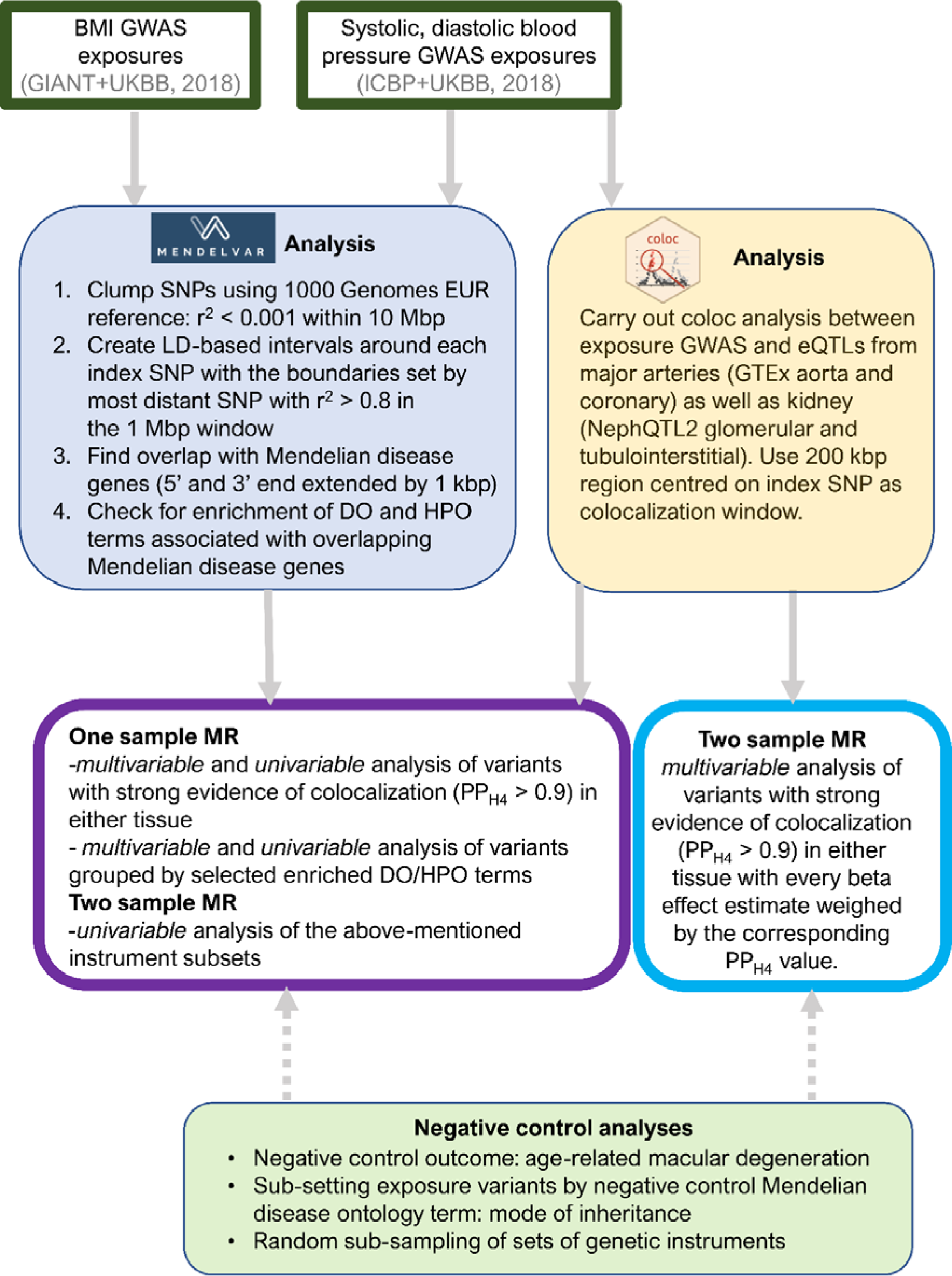
An overview of this study’s workflow for Mendelian disease gene-derived partitioning of genetic instruments and colocalization-derived partitioning of genetic instruments with the aim of investigating pathway-specific effect of blood pressure and body mass index on cardiometabolic traits.

## Materials and Methods

### Exposure and outcome GWAS datasets

Our exposure datasets consisted of systolic (SBP) and diastolic blood pressure (DBP) GWAS produced by ICBP in 2018^27^ and body mass index (BMI) produced by the GIANT consortium in 2018^40^ (**Supplementary Table 1**). These large studies, comprising ∼1 million and ∼0.7 million individuals of European ancestry, respectively, were highly powered and returned a number of top variants (∼900) implicated in a variety of biological processes in each GWAS.

Our outcome GWAS datasets included common cardiometabolic diseases: atrial fibrillation (AF)^41^, heart failure (HF)^42^, coronary heart disease (CHD)^43^, myocardial infarction (MI)^43^, stroke^44^, and type 2 diabetes (T2D, see **Supplementary Table 1** for details). GWAS for continuous measurements of cardiac function included: left ventricular end-diastolic volume (LVEDV), left ventricular end-systolic volume (LVESV), left ventricular ejection fraction (LVEF), and left ventricular stroke volume (SV).

As a negative control, we selected an outcome whose incidence is likely not to be causally impacted by BMI or BP as indicated by a previous MR study^45^: age-related macular degeneration (AMD)^46^.

All the GWAS summary statistics, with the exception of left ventricular function traits (downloaded from https://personal.broadinstitute.org/ryank/) and early-onset AMD (downloaded from https://homepages.uni-regensburg.de/~wit59712/earlyamd/winkler_et_al_earlyamd_meta.gz) were accessed via Open GWAS^47^.

We note that we used exposure and outcome GWAS datasets with non-overlapping participants whenever feasible, to limit bias^48^, however a significant proportion of individuals in the AF and early-onset AMD (∼20-40%), as well as in the left ventricular function GWASes were obtained based on UK Biobank individuals, who are also included in our exposure GWAS for BMI and BP.

### Genetic instrument selection

BMI and BP GWAS summary statistics were obtained in GWAS-VCF^49^ format from the OpenGWAS platform, and were subsequently converted to the TwoSampleMR^50^ package format using the gwasvcf_to_TwoSampleMR function from the gwasglue (https://mrcieu.github.io/gwasglue/) R package. In order to identify independent genetic instruments for each exposure, we first filtered SNPs that showed strong association at a genome-wide significance level (p-value < 5 × 10^-8^). Next, the SNPs were clumped using ld_clump wrapper for plink ver 1.943^51^ from the ieugwasr R package (https://mrcieu.github.io/ieugwasr) to ensure that linkage disequilibrium (LD) as measured by r^2^ was < 0.001 within 10 Mbp in 1000 Genomes^52^ European panel. We sometimes used SNP proxies showing high genetic correlation (r^2^ > 0.8) in the instances when the chosen SNP was missing in the outcome dataset.

### Assignment of genetic instruments to pathways: Mendelian disease

Having obtained 887 independent SNP instruments for BMI, 914 for diastolic BP and 863 for systolic BP, we used MendelVar^19^ to partition the variants into subsets enriched for Mendelian disease categories. Briefly, we used the MendelVar^19^ pipeline to generate LD-based genomic intervals around each input SNP (delimited by most distant SNP with minimum r^2^ = 0.8 within 1 Mbp in either direction). Then, we checked for enrichment of phenotype ontology terms linked to Mendelian disease genes (defined as coding region with 1000 bp 5’ and 3’ flanking regions) present within the interval using the INRICH^53^ software contained in the MendelVar platform.

### Assignment of genetic instruments to tissues: colocalisation

We followed the method described in Leyden et al. (2022)^11^ to assign SNPs to subsets with eQTL colocalisation evidence in at least one of the two chosen tissue types (kidney and vasculature). For BP traits, we used cis-eQTLs in kidney (NephQTL2^54^ tubulointerstitial *n*=311 and glomerular *n*=240) and arteries (GTEx 8^55^ aorta *n*=387 and coronary *n*=212); similar sample sizes across tissues should result in comparable power. We used intervals of +/- 100 kbp centred on each exposure SNP for colocalisation in the coloc R software^56^, and a stringent posterior probability H4 (PP_H4_, hypothesis 4: shared causal variant between exposure GWAS and cis-eQTL dataset) threshold of 0.9 to partition SNPs into coloc-based tissues.

### Enrichment analysis within subsets

We used ToppFun from ToppGene Suite^57^ and over-representation module in ConsensusPathDB^58^ (both with default settings) to test for global enrichment of functional terms in gene subsets identified by MendelVar and coloc harnessing popular ontologies, such as GO^59^, Reactome^60^ and KEGG^61^.

### One sample Mendelian Randomization analyses

Our methodology for one-sample MR analyses followed the protocol described in Leyden et al. (2022)^11^ and utilized individual-level data from the UK Biobank ^62^(Application number: 81499). This involved the creation of a genetic risk score (GRS), factoring in all SNPs or coloc/Mendelian SNP subsets, weighted according to their respective effect sizes. The sample comprised 334,398 unrelated people of European descent, and was established after excluding participants with withdrawn consent, genetically related, or those who did not cluster with “white European” group based on K-means clustering (K=4).

Subsequently, we used all exposure (BMI, DBP, SBP) SNPs to derive MR estimates employing either linear or logistic regression using AER and mass R packages. We analysed all available outcomes (**Supplementary Table 2**: AF, HF, CHD, MI, T2D, stroke, AMD, LVEDV, LVESV, LVEF, SV) and adjusted for variables such as age, sex, the leading 10 principal components, and a binary marker for genotype chips. A rank-based inverse normal transformation was applied to continuous cardiac outcomes (LVEDV, LVESV, LVEF, SV) before analysis. We carried out the univariable analysis using each coloc or Mendelian SNP GRS as exposure in turn.

Finally, we deployed multivariable models which incorporated either the two coloc- or two Mendelian-based SNP subset GRS to quantify the direct effect of each subset on the outcome.

To estimate the type 1 error rate due to the fraction of shared individuals between our GRS construction GWAS datasets and the cases and controls in the UK Biobank sample, we used the ‘‘sample overlap” web app (https://sb452.shinyapps.io/overlap/).The results indicated that, based on the strong F-statistics of our instruments, the overlap of samples is unlikely to cause significant bias in our analyses (type 1 error rate < 0.05).

### Sensitivity analyses

We also used GWAS exposure and outcome datasets described above in two-sample MR analysis. Firstly, we carried out univariable MR analysis, using all top SNPs for a given exposure and then Mendelian or coloc SNP subsets separately. Secondly, for coloc SNP subsets we ran multivariable MR including both tissue subsets to obtain mutually adjusted “independent” effect as per Leyden et al. (2022). In this analysis, each instrument’s effect size is weighted by coloc’s PP_H4_ in its target tissue. We were not able to carry out multivariable MR for Mendelian SNP subsets due to lack of a suitable metric by which to scale individual variants’ effects. We chose the “TwoSampleMR”^50^ R package for the standard inverse-variance weighted, mode-weighted, median-weighted and MR-Egger MR analyses as well as multivariable MR analysis. We then calculated instrument’s strength (F-statistics, R^2^) and heterogeneity (Cochran’s Q and I^2^)^63^.

In addition to including a negative control for the outcome (AMD), we also attempted to provide a negative control SNP partitioning in the Mendelian disease context. We subset the exposure SNPs by a feature which was not expected to biologically influence the outcome (mode of inheritance: autosomal dominant or recessive) and which was not significantly enriched in any Mendelian blood pressure SNP set. We did not run this control example for BMI exposure, since we did find enrichment of autosomal dominant disease-assigned SNPs among mental health disease SNPs in the dataset, mostly driven by intellectual development disorders (χ^2^=10.3, N=71, p- value=0.001, **Supplementary Table 3**).

### Random sampling of SNP subsets

Finally, we decided to empirically determine how often the difference in MR estimates between randomly drawn SNP subsets equals or exceeds the one observed for Mendelian or coloc-partitioned instruments. When simulating subsets for comparison with Mendelian disease-partitioned instruments, we drew random *n_1_*SNPs without replacement and then separately *n_2_* SNPs without replacement (where *n_1_* and *n_2_* correspond to the number of SNPs in the original SNP subsets) to represent two SNP subsets which can randomly overlap. For coloc, the procedure for drawing SNPs was modified to randomise the association between SNP, tissue and colocalisation probability. In that case, we first randomly permuted all PP_H4_ values across both tissues and variants, following which we extracted SNPs with PP_H4_ values above the chosen threshold (0.9 in our analysis, and 0.8 in replication of Leyden et al. (2022), see below) to be used as two random SNP subsets. We then ran equivalent 1 sample and 2 sample MR analyses as for the “true” coloc and Mendelian SNP subsets. The entire procedure was repeated 1,000 times per each exposure-outcome and analysis type combination.

### Replication of Leyden et al. (2022)

We re-analysed the Leyden et. al (2022)^11^ dataset for select outcomes to compare their MR results using BMI exposures assigned by coloc to the adipose (86 SNPs) and brain (140 SNPs) tissues with our Mendelian disease-partitioned approach. We also established the extent to which the difference between coloc-partitioned SNP subsets in the MR analyses could emerge by chance, using random instrument re- sampling as described above.

## Results

### Assignment of variants to pathways - Mendelian

Using MendelVar, we assessed overrepresentation of Mendelian disease genes (and their symptoms) in strong LD with blood pressure GWAS top loci (**Figure 1**). In particular, we were interested in investigating enrichment of Mendelian phenotypes related to kidney as opposed to vasculature. The two most enriched terms found were “abnormal renal morphology” (DBP: 90 SNPs, p-value=8x10^-5^; SBP: 84 SNPs, p-value=2.8x10^-4^, **Supplementary Table 4-7**) and “abnormal blood vessel morphology” (DBP: 79 SNPs, p-value=1.2x10^-4^; SBP: 83 SNPs, p-value=2x10^-5^) from the Human Phenotype Ontology (HPO)^64^. The instruments in the “renal” and “vessel” subsets showed substantial overlap: 37 for diastolic SNPs and (p-value= 1.2x10^-4^) 42 for systolic SNPs (p-value=2x10^-6^, **Supplementary** Figure 1A-B).

We then used independent ontologies (featured in ConsensusPathDB^58^ and ToppGen^57^ resources) not related to Mendelian disease to check if they provide orthogonal enrichment evidence for the role of the assigned genes in a given pathway or tissue (**Supplementary Tables Enrichment (STE)** and **Supplementary Results**). We found that kidney-related terms were significantly enriched in the “renal” gene set (**Supplementary Dataset STE 1-4**), e.g. renal system development (Gene Ontology^65^, DBP q-value: 2.6x10^-9^; SBP q-value: 4.7x10^-6^). However, we also observed a significant overrepresentation of gene sets related to metabolism, hormonal regulation, type 2 diabetes and cancer. Our “vessel” gene set was found to contain a strong overrepresentation of cardiovascular terms across many ontologies (**Supplementary Dataset STE 5-8**): e.g. blood vessel development (Gene Ontology, DBP q-value: 7.7x10^-6^; SBP q-value: 4.8x10^-10^). To a lesser extent, we also found enrichment of kidney-related terms (*renin secretion*, *EPO signalling pathway*) in the “vessel” gene set and cardiovascular-related terms (*blood vessel development*, *heart development*) in the “renal” set which is not unexpected given substantial overlap of “renal” and “vessel” SNPs.

For BMI, we used the Alliance of Genome Resources slim (28 general disease types) version of Disease Ontology^66^ where the top most enriched categories were “disease of metabolism” (45 SNPs, p-value=8.8x10^-4^, **Supplementary Tables 8-9**) and “disease of mental health”/”developmental disorder of mental health” (39 SNPs, p-value=4.1x10^-3^). Choosing this ontology allowed us to contrast the contribution of variants related to metabolism and brain function, which have previously shown distinct patterns across disease outcomes^11^. The “metabolic” and “mental health” SNP subsets were largely disjoint, with only 6 shared SNPs (p-value=0.71, **Supplementary** Figure 1 **C**). The good separation of “mental health” and “metabolic” gene sets was reflected in the top terms enriched in non-Mendelian disease functional ontologies (**Supplementary Dataset STE 9-12**). The most enriched terms in the “mental health” set related to the brain, in particular synaptic signalling, e.g. neuronal system (Reactome^67^, q-value: 1x10^-3^). Reassuringly, the strongest enrichment of terms in the “metabolic” gene set related to metabolism and type 2 diabetes, e.g. metabolism (Reactome, q-value= 5x10^-11^).

### Assignment of variants to tissues – coloc

We then applied the previously proposed^11^ approach with colocalisation used to assign lead blood pressure SNPs to tissues as an alternative method to investigate the difference in SNP sets with roles primarily in kidney or vasculature (**Supplementary Tables 10-11**). Use of high minimum posterior probability of colocalisation threshold (PP_H4_ > 0.9) for blood pressure top loci resulted in assignment of 117 SNPs to the aorta or coronary (referred to as “artery” collectively) tissue for DBP and 132 for SBP. In kidney, 87 SNPs colocalised in the glomerular or tubulointerstitial tissues (referred to as “nephro” collectively) for DBP and 77 for SBP (**Supplementary Tables 12-13**). Among “artery” SNPs, the majority of loci colocalised in the aorta (108 for SBP, 126 for DBP; **Supplementary** Figure 1 D-E), and the loci shared between the aorta and coronary artery amounted to approximately one-third of all. Among “nephro” SNPs, tubulointerstitial tissue dominated with 65 and 64 SNPs for DBP and SBP (**Supplementary** Figure 1 D-E), but less sharing between the two tissue types was found than in the “artery” subsets – one-quarter and one-fifth was common to both tubulointerstitial and glomerular in the DBP and SBP variant sets, respectively. Comparison of “artery” and “nephro” SNP sets revealed some overlap, with 36 SNPs shared in DBP (p-value<1x10^-6^) and 42 SNPs shared in SBP (p-value<1x10^-6^).

Coloc-derived assignment showed limited alignment with functional pathways relevant to each tissue (**Supplementary Dataset STE 13-20**), with very few weakly enriched terms found overall: 4 for “artery” SNPs in DBP (hypertrophic cardiomyopathy, ACE inhibitor pathway, metabolism of lipids, mitochondrial electron transport chain; q-value=0.029-0.047), 3 for “nephro” SNPs in DBP (O-glycosylation of TSR domain-containing proteins, aquaporin-mediated transport, transport of small molecules; q-value=0.024-0.047) and 3 cardiomyopathy terms were enriched for among “nephro” SBP genes (q-value=0.017-0.024).

BMI colocalization results were obtained from Leyden et al. (2022). Among those, 140 SNPs were assigned to the brain and 86 to the adipose tissue, with 43 overlapping (p-value<1x10^-6^). Enrichment of genes with colocalisation evidence for BMI in adipose and brain tissue sets showed limited overlap with biologically relevant terms, especially for the brain (**Supplementary Dataset STE 21-24**); this replicated analyses previously described in Leyden et al. (2022).

### Comparison of Mendelian- and coloc- derived SNP sets

Finally, comparison of SNP sets derived using Mendelian- and coloc- assignment methods showed that they are largely distinct and could potentially offer orthogonal evidence (**Supplementary** Figure 1 F-G). Altogether, 33 DBP (p-value=0.35) and 34 SBP (p-value=0.41) SNPs were found in both Mendelian and coloc-based SNP subsets, as opposed to 234 and 234 non-shared SNPs, accordingly. Similar observations were made for BMI (**Supplementary** Figure 1 H-I): only 5 out of the 45 “mental health” Mendelian SNPs were shared with coloc groupings as assigned previously by the Leyden et al. (2022) study – 4 with “brain” and 1 with both “adipose”/”brain” (adipose p-value=0.98, brain p-value=0.77). For “metabolic” Mendelian SNPs higher overlap was found: 17 out of 39 SNPs were shared with coloc subsets (p-value=0.02): 3 SNPs with “adipose“, 5 SNPs with both “adipose“/“brain“ and, unexpectedly, 9 SNPs with “brain“ colocalising SNPs (adipose p-value=0.06, brain p-value=0.006).

### Pathway-based Mendelian Randomization analyses for blood pressure

Having established the Mendelian disease- and coloc-derived SNP sets for blood pressure, we then proceeded to use them as exposures in one sample MR studies carried out in individuals of European ancestry in the UK Biobank. We focussed on selected cardiometabolic outcomes (**Supplementary Table 2**), as they have been firmly established to be causally influenced by blood pressure^22,68–70^ and were previously evaluated using the coloc-based approach with respect to BMI by Leyden et al. (2022).

We confirmed that DBP and SBP have a strong causal relationship with coronary heart disease (**Figure 2, Supplementary Table 14**): OR_95%CI_=1.09-1.11 and OR_95%CI_=1.06-1.07, respectively. Selecting Mendelian-based SNP subsets (“renal” and “vessel”) and adjusting for shared effect in multivariable MR analysis (**Supplementary Table 15**) containing both subsets indicates that the “vessel” subset is driving the positive effect of DBP and SBP on CHD (DBP: OR=1.17, OR_95%CI_=1.14-1.21, p-value=1.5x10^-28^; SBP: OR=1.13, OR_95%CI_=1.11-1.15, p-value=2x10^-43^), relative to “renal” subset (DBP: OR=1.04, OR_95%CI_=1.01-1.07, p-value=6.2x10^-3^; SBP: OR=1.01, OR_95%CI_=0.99-1.02, p-value=1.9 x10^-2^), with 95% confidence intervals of the two subsets distinct and non-overlapping.

**Figure 2.**
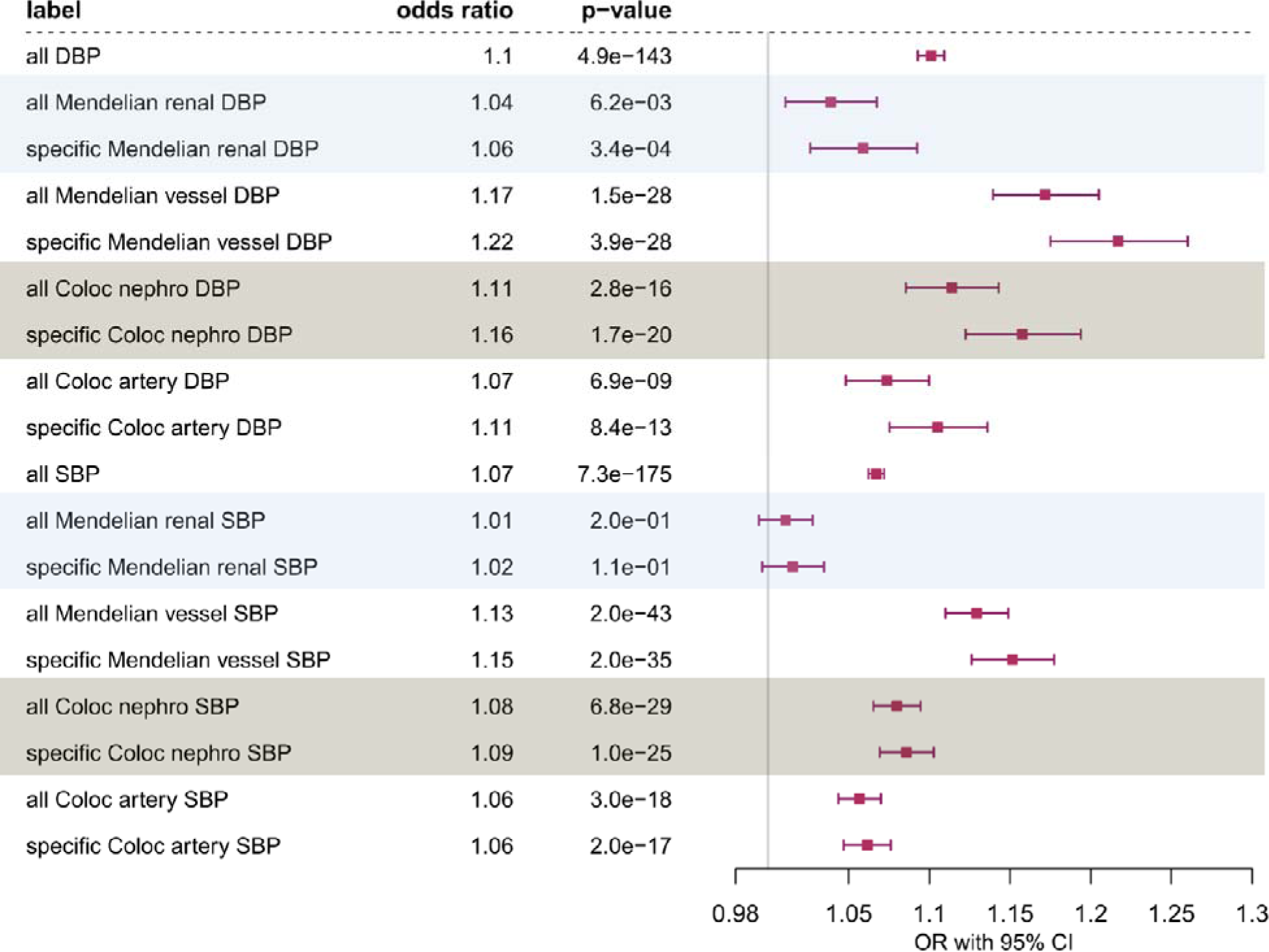
Coronary heart disease: one sample Multivariable Mendelian Randomization analysis of the effect of diastolic blood pressure (DBP) and systolic blood pressure (SBP) on CHD using all SNPs, all/specific Mendelian disease partitioned (disease with abnormalities in the *renal* or blood *vessel* system) genetic instruments and all/specific Coloc partitioned (*nephro* – kidney tissues: glomerular and tubulointerstitial, *artery* – aorta and coronary artery tissues) instruments. Effect sizes are scaled to per one SD change in blood pressure.

However, comparison with coloc-based subsets (**Supplementary Table 16**) revealed the opposite trend with the kidney “nephro” subset (DBP: OR=1.11, OR_95%CI_=1.08-1.14, p-value=2.8x10^-16^; SBP: OR=1.08, OR_95%CI_=1.06-1.09, p-value=6.8x10^-29^) showing a stronger effect size than the “artery” SNP subset in the MVMR analysis (DBP: OR=1.07, OR_95%CI_=1.05-1.10, p-value=6.8x10^-9^; SBP:

OR=1.06, OR_95%CI_=1.04-1.07, p-value=3x10^-18^). All of the coloc exposures provided evidence of causal effect on CHD for SBP and DBP, unlike Mendelian “renal” SNP subset for SBP which does not seem to causally influence CHD (p*-*value=0.19).

Limiting exposures to SNPs specific to each subset resulted in similar causal estimates as when using all SNPs. We also show similar results for myocardial infarction (**Supplementary** Figure 2) which is closely genetically correlated to CHD (**Supplementary** Figure 3). Point estimates for MI display greater uncertainty and while the difference between Mendelian disease-partitioned exposures persists, it was not apparent for “artery” and “nephro” coloc-based subsets.

On the other hand, coloc and Mendelian-based SNP subsets showed consistent direction of effect for an important cardiac function outcome, left ventricular stroke volume (SV, **Figure 3**). Analyses including all SNPs show reduction in stroke volume on increase in DBP (beta=-0.015, beta_95%CI_=(-0.021,-0.008), p-value=2.9x10^-6^), but the opposite for SBP (beta=0.006, beta_95%CI_=(0.002,0.01), p-value=2.7x10^-3^). There is limited support for “renal” (beta_95%CI_=(-0.013,0.034) and “nephro” (beta_95%CI_=- 0.011,0.034) diastolic BP exposures being causally associated with SV. We find that it is the “vessel” (beta=-0.052, beta_95%CI_=(-0.076,-0.028), p-value=2.8x10^-5^) and “artery” diastolic BP (beta=-0.03, beta_95%CI_=(-0.051,-0.009), p-value=5.6x10^-3^) exposures that drive the negative effect of DBP on stroke volume. Mendelian “vessel” SNPs also have a certain negative effect on systolic BP (beta_95%CI_=(-0.034,- 0.004)) while the “renal” SNPs contribute to the overall positive effect of SBP (beta_95%CI_=(0.0003,0.0289)), however there is no clear distinction for coloc- partitioned exposures.

**Figure 3.**
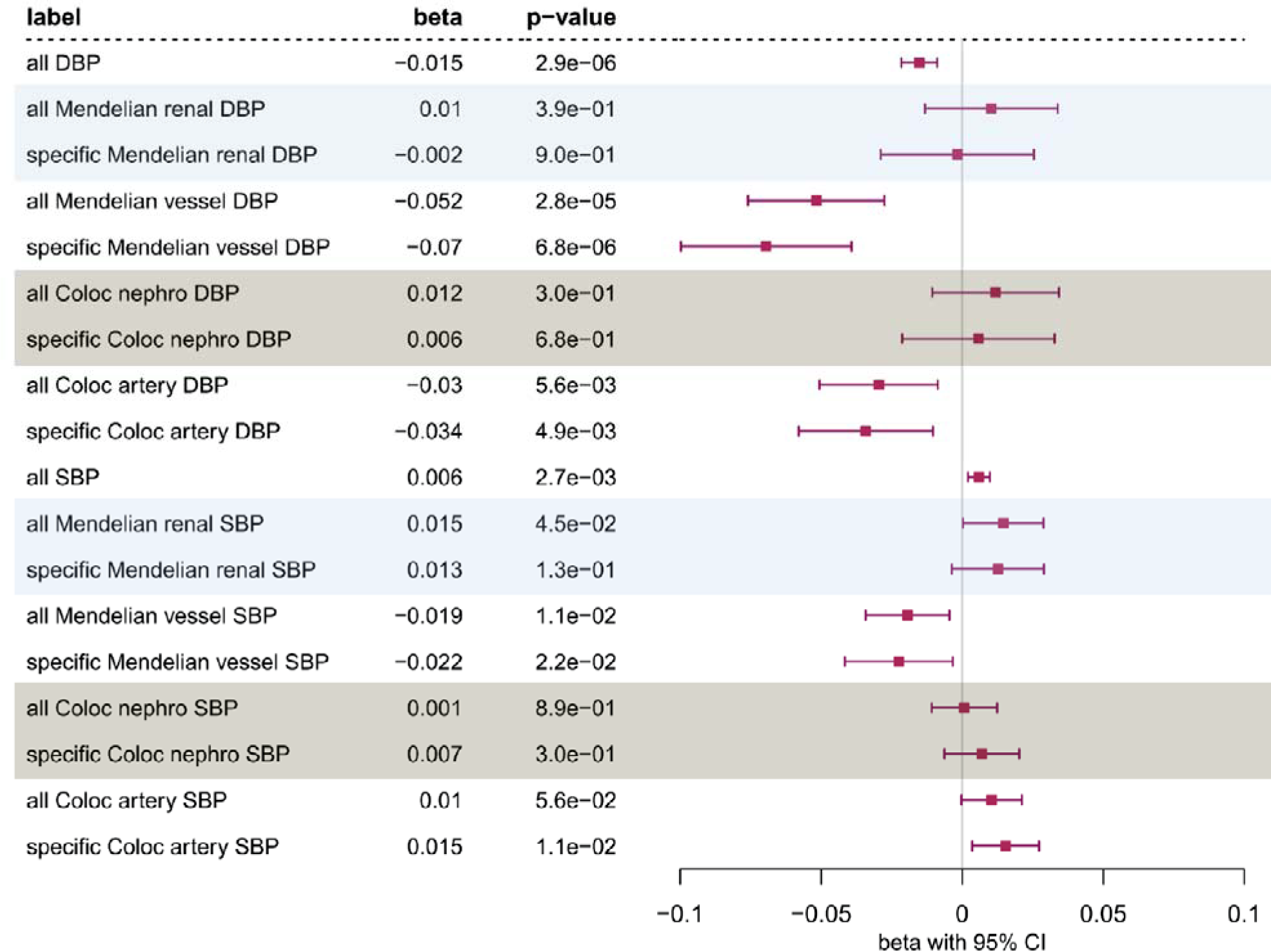
Stroke volume: one sample Multivariable Mendelian Randomization analysis of the effect of diastolic blood pressure (DBP) and systolic blood pressure (SBP) on stroke volume (SV) using all SNPs, all/specific Mendelian disease partitioned (disease with abnormalities in the *renal* or blood *vessel* system) genetic instruments and all/specific Coloc partitioned (*nephro* – kidney tissues: glomerular and tubulointerstitial, *artery* – aorta and coronary artery tissues) instruments. Effect sizes are scaled to per one SD change in blood pressure.

Dissecting the effect of various SNP subsets on type 2 diabetes reveals a heterogenous landscape (**Figure 4**). We first confirmed the increase in the odds of T2D associated with elevated blood pressure (DBP OR_95%CI_=1.02-1.04; SBP OR_95%CI_=1.03-1.04). Next, we found that Mendelian-disease partitioned subsets show comparable effect on the risk of T2D for DBP, but the 95% confidence intervals are wide. Whilst “vessel” SBP exposure produces higher odds of T2D (OR=1.07, OR_95%CI_=1.05-1.09, p-value=1.5x10^-12^) than “renal” SBP exposure (OR=1.03, OR_95%CI_=1.01-1.05, p-value=4.2x10^-4^), effect size estimates are partially overlapping. In agreement with this pattern, coloc-based “artery” SNPs have a positive causal effect on the risk of T2D for both DBP (OR=1.08, OR_95%CI_=1.05-1.11, p-value=4.3x10^-8^) and SBP (OR=1.06, OR_95%CI_=1.04-1.07, p-value=8.6x10^-17^), while null effect was found for “nephro” SNPs (DBP OR_95%CI_=0.97-1.03; SBP OR_95%CI_=0.99-1.02).

**Figure 4.**
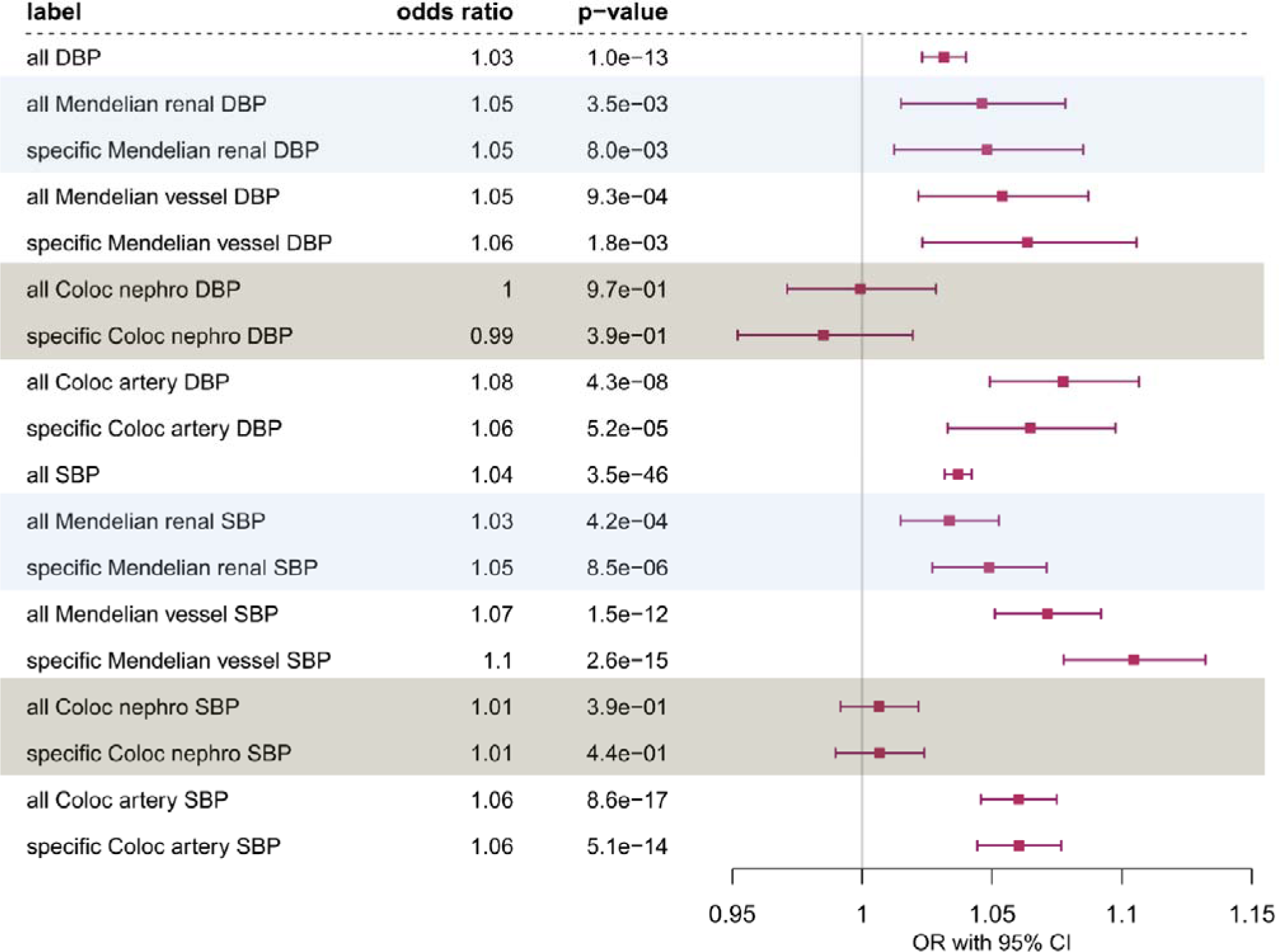
Type 2 diabetes: one sample Multivariable Mendelian Randomization analysis of the effect of diastolic blood pressure (DBP) and systolic blood pressure (SBP) on T2D using all SNPs, all/specific Mendelian disease partitioned (disease with abnormalities in the *renal* or blood *vessel* system) genetic instruments and all/specific Coloc partitioned (*nephro* – kidney tissues: glomerular and tubulointerstitial, *artery* – aorta and coronary artery tissues) instruments. Effect sizes are scaled to per one SD change in blood pressure.

Repeating the analysis above and analysing each exposure subset independently using one sample MR univariable framework (**Supplementary Table 15-16**) produces similar patterns between SNP subsets, with any differences in effect estimates accentuated further in MVMR analyses.

### Sensitivity analyses for blood pressure

We used summary-level statistics based on independent samples for exposure and outcome to see if two-sample MR returns results in agreement with those obtained in one-sample MR (**Supplementary Table 17- 19**). In general, we discovered that two-sample analyses had less power to detect any differences in effect between SNP subsets, but direction of effect was consistent with that observed in one-sample MR. The one exception was a more pronounced causal effect of “nephro” SBP exposure on myocardial infarction (OR=1.04, OR_95%CI_=1.02-1.06, p-value=2.3x10^-4^) relative to “artery” (OR=1, OR_95%CI_=0.98-1.02, p-value=1), which corresponds to similarly dominant role of “nephro” SBP SNPs in one-sample MVMR for the related outcome – CHD.

We found that mean absolute effect sizes tend not to differ significantly between all SNPs and Mendelian-/coloc-based exposure subsets (**Supplementary Table 20**). Among all SNP subsets, mean F-statistics was > 60 suggesting low risk of weak instrument bias.

We then proceeded to evaluate changes in the average heterogeneity (*Q_het_*=*Q/(Q_df_- _1_*)) as we expected that it should decrease within tissue- or pathway-based SNP subsets in comparison to all SNPs representing a variety of pleiotropic processes. However, we did not detect a downward trend in average heterogeneity which instead varied in unexpected ways. For example, we found *Q_het_* to be lower among Mendelian-partitioned and coloc-partitioned exposures relative to all DBP SNPs (**Supplementary Table 21-23**). However, for SBP Mendelian “renal” and coloc “artery” SNP subsets showed higher *Q_het_* than all SBP SNPs, while Mendelian “vessel” and coloc “nephro” SNP subsets were the opposite.

Next, we repeated our analyses using a negative outcome phenotype: age-related macular degeneration^45^. The one-sample and two-sample MR results (**Supplementary Table 15-16, 18-19**) did not indicate strong evidence of causal effect for our coloc- or Mendelian- based SNP subsets, in line with our expectations. We did find one weak non-null result, however, only at a nominal level (**Supplementary** Figure 4) for SNPs specific to the Mendelian “renal” subset in DBP and SBP (DBP OR=1.07, OR_95%CI_=1-1.14, p-value=0.03; SBP OR=1.04, OR_95%CI_=1.00–1.08, p-value=0.03).

Negative control partitioning of exposures by main modes of Mendelian disease inheritance (autosomal “dominant” or “recessive”) showed no association with the Mendelian “renal”-”vessel” subdivision (**Supplementary** Figure 5 **A, B**) for DBP (p-value=0.13) and SBP (p-value=0.4, **Supplementary Table 3**). We also assumed that this negative control feature should not be associated with biologically meaningful pathway- or tissue- based partitioning of exposure SNPs, which may not hold. We did indeed detect a substantial difference between “dominant” and “recessive” SNPs sets when focussing on CHD as outcome in one sample MR analysis (**Supplementary** Figure 6). For instance, all “dominant” diastolic BP SNPs had a higher effect on CHD (OR=1.12, OR_95%CI_=1.09-1.14, p-value=3.9x10^-23^) than all “recessive” SNPs (OR=1.08, OR_95%CI_=1.05-1.10, p-value=1.8x 0^-11^) (**Supplementary Table 24**). However, when limiting ourselves to exposure SNPs “specific” to each subset, the opposite conclusion was obtained with higher effect size seen for “recessive” SNPs (OR=1.22, OR_95%CI_=1.17-1.26, p-value=3.9x10^-28^) than “dominant” (OR=1.06, OR_95%CI_=1.03-1.09, p-value=3.4x10^-4^). No meaningful difference between the SNP subsets was observed in two-sample MR results (**Supplementary Table 25, 26**). In addition, much less variation across SNP subsets was found for the T2D outcome (**Supplementary** Figure 7), with the exception of one outlier: specific “recessive” SBP SNP subset.

### Pathway-based Mendelian Randomization analyses for body mass index

We found that the Mendelian disease-partitioned BMI SNPs showed the largest difference between “mental health” and “metabolic” SNPs for atrial fibrillation. All BMI SNPs confirmed a moderate effect on AF (OR=1.05, OR_95%CI_=1.04-1.06, p-value=4.6x10^-55^; **Supplementary Table 27**). This effect size was then matched by “mental health” SNPs in one sample multivariable MR analysis (OR=1.05, OR_95%CI_=1.02=1.08, p-value=5.6x10^-4^, **Supplementary Table 28**) but a larger effect size was found for “metabolic” exposures (OR=1.10, OR_95%CI_=1.07-1.13, p-value=3.3x10^-10^). Similar magnitude of differences between “all”, “metabolic” and “mental health” exposures was obtained in 1 sample univariable analyses (**Supplementary Table 28**) and 2 sample MR analyses (**Supplementary Table 29- 30**). This is in contrast to coloc-based SNP stratification carried out by Leyden et al. (2022) and replicated here (**Supplementary Table 31-34**), where the “brain” SNP subset was found to increase the risk of AF more strongly (1 sample MVMR OR=1.04, OR_95%CI_=1.02-1.06, p-value=6.4x10^-6^, **Supplementary Table 32**), than the “adipose” SNP subset (1 sample MVMR OR=1.02, OR_95%CI_=1-1.04, p-value=0.08) but the effect size difference was relatively attenuated.

### Sensitivity analyses for body mass index

Average heterogeneity (*Q_het_*) of “mental health” and “metabolic” BMI SNPs for AF was close to average heterogeneity for all BMI SNPs (**Supplementary Table 35-36**) – 2.5, 2.05, 2,19, respectively. We also did not uncover evidence for systematically reduced average heterogeneity among SNP subsets across other outcomes.

Results from negative control outcome (AMD) indicated that “mental health” and “metabolic” SNP subsets analysed independently in 1 sample/2 sample univariable MR or in conjunction with each other (1 sample multivariable MR) show no evidence for causal role, as per expectations (**Supplementary Table 28, 30**).

### Random sampling of SNP subsets

Overall, we noticed that one sample MVMR results showed a variety of effect size differences between exposure SNP subsets, ranging from negligible (e.g. diastolic BP Mendelian partitioning with respect to T2D), medium with overlapping confidence intervals (systolic BP Mendelian partitioning with respect to T2D) to totally distinct (systolic BP Mendelian partitioning with respect to CHD). Conclusions from our negative control outcomes and exposures were unclear as we sometimes found substantial differences in effects between Mendelian and coloc exposure subsets which appeared not to satisfy our null expectations. Therefore, we found it difficult to establish whether any differences detected were likely to be both biologically meaningful and not driven by chance assortment. To address this issue, we employed a simulation technique, where we re-ran our analysis pipeline 1000 times for each exposure-outcome pair using 2 subsets of SNP sampled randomly from across all the exposure SNPs. This allowed us to quantify the frequency of the absolute differences in effect sizes such as observed in our MR analysis (or greater) relative to background (in the process obtaining two-tailed p-value with the floor value of 10^-3^), and empirically derive 95% confidence intervals of the difference (**Supplementary Table 37**).

Figure 5 provides an overview of this sensitivity approach run for all the exposures (BMI, SBP, DBP), SNP subsets (Mendelian, coloc), MR methods (1 sample or 2 sample) and MR models (univariable, multivariable). Across all the MR analyses with blood pressure as exposure, we find strong evidence for the differential effect of Mendelian “renal” and “vessel” exposures on CHD (p-value range: 0.001-0.042) along with MI. In addition, coloc “artery” and “nephro” systolic BP subsets show a strong difference of effect on T2D, CHD and MI (p-value range: 0.007-0.042) but the latter two only in the two-sample MR setting. Significant evidence based on only 1 sample MVMR was found among coloc-based SNP subsets for diastolic BP and T2D, as well as BMI and CHD/MI. Both one-sample and two-sample MVMR analyses support a differential effect of coloc-based “adipose” and “brain” SNPs on BMI (p-value=0.001-0.013).

**Figure 5.**
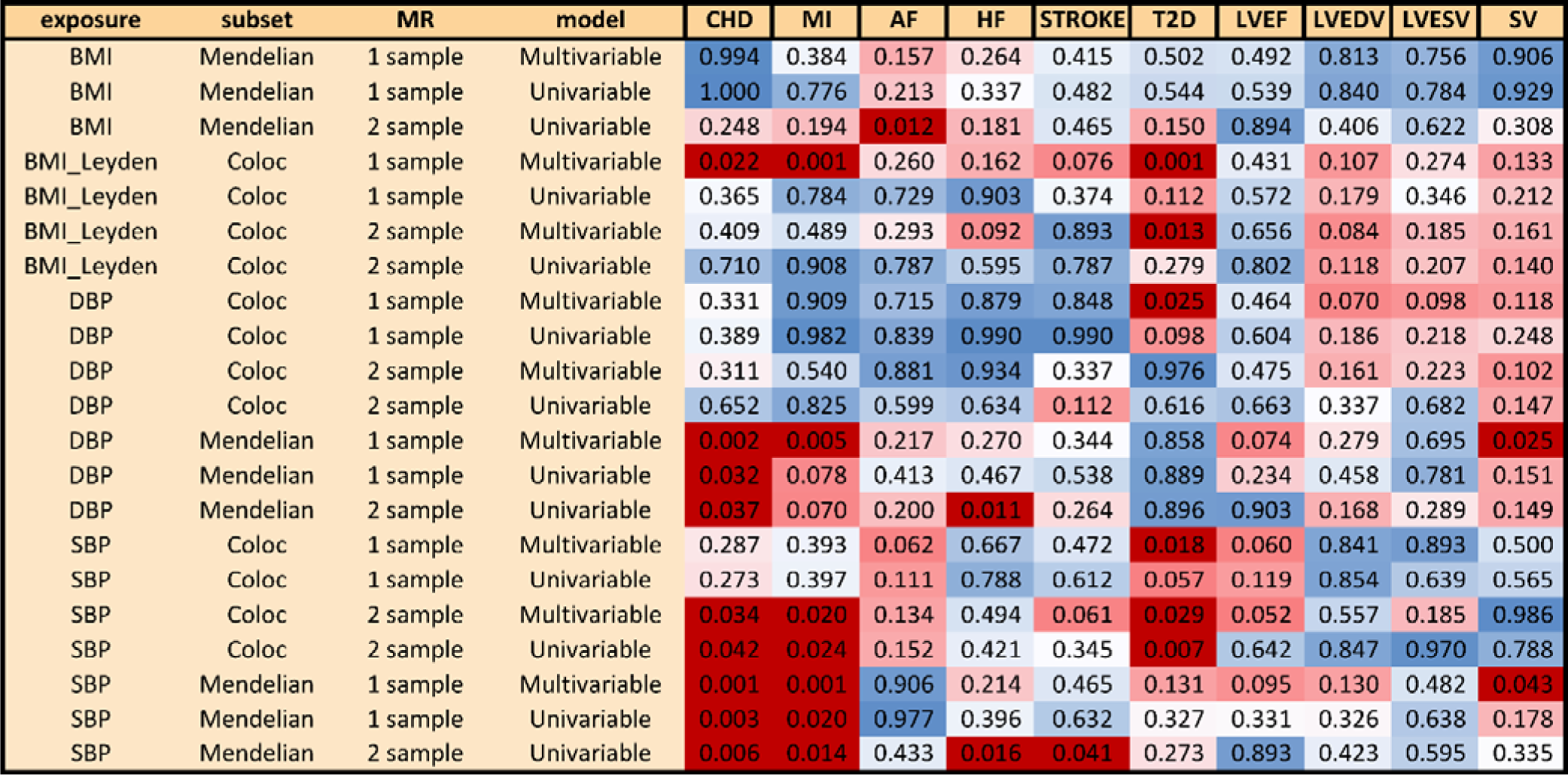
Matrix of empirically-derived (1,000 replicates) p-values for distribution of effect size differences between Mendelian disease-partitioned and coloc-partitioned instruments in 1 sample and 2 sample MR setting (Univariable and Multivariable) using body mass index (BMI) and blood pressure – systolic (SBP) and diastolic (DBP) as exposures and cardiometabolic traits as outcomes (AF, CHD, MI, HF, stroke, T2D, LDVEDV, LVEF, LVESV, SV). P-values < 0.05 are highlighted in dark red.

We also find weaker evidence for a difference between “renal” and “vessel” blood pressure subsets with respect to the stroke volume outcome (1 sample MVMR SBP p-value= 0.043, DBP p-value=0.025). There was little evidence against the null hypothesis (of chance SNP assortment) for any of the BMI or BP pathway-based results with respect to atrial fibrillation, heart failure, stroke, LVEDV, LVESV or LVEF, with the exception of the following two-sample univariable MR analyses involving Mendelian disease-partitioned instruments: BMI effect on atrial fibrillation (p-value=0.012), systolic and diastolic BP effect on heart failure (p-value < 0.02).

## Discussion

In this investigation, we employed MR to dissect the causal associations between blood pressure or body mass index and cardiometabolic traits, leveraging biological pathway information. To do so, we classified genetic instruments into Mendelian disease categories with MendelVar or assigned SNPs to tissues through colocalization. Our findings underscore the significance of renal and vascular genes in blood pressure-related conditions, revealing distinct sets of SNPs from Mendelian and colocalization methods. Notably, while both Mendelian and colocalisation-based SNP sets implicated diastolic and systolic BP in coronary heart disease, they emphasized different physiological routes – vessel and kidney, respectively. We report similarly contrasting results for the effect of BMI on atrial fibrillation, when comparing estimates using Mendelian- (“metabolic”-led) and coloc- (“brain”-led) derived instrument subsets. Sensitivity analyses emphasized the consistent direction of effects in one-sample and two-sample MR (univariable and multivariable) but showed some variability in magnitude. We also conducted simulations to assess the probability that differences in causal estimates between SNP subsets arose by chance, and these confirm the validity of interpretation of our main findings.

In the main results, we presented examples of representative cardiac traits – CHD and SV, which as shown in Supplementary Figure 2 are highly genetically correlated with MI and LVEDV/LVESV, respectively. As expected, and demonstrated in Figure 5, magnitude of effect size difference between Mendelian disease- and coloc- partitioned SNP subsets for those related exposures is highly congruent. For ventricular function traits, we only focus on one-sample MR as two-sample results can be biased due to sample overlap of UK Biobank individuals with our exposure sample. The interesting outlier effect of increased importance of “metabolic” SNPs relative to “mental health” observed for atrial fibrillation in two-sample univariable MR could be also potentially affected by the sample overlap bias.

A certain limitation to the current Mendelian disease-partitioning approach is the inability to model the subsets jointly using the multivariable approach in a two-sample MR setting. Altogether, we find a comparable number of results (14) with empirically determined significant differences (p-value < 0.05) in one-sample (univariable: 3, multivariable: 11) versus two-sample MR (univariable: 10, multivariable: 4), with univariable and multivariable results mostly agreeing in the direction and magnitude of effect for the same SNP subsets. In some cases, we found reassuring agreement across all MR types and model specifications: Mendelian subsets for diastolic BP versus CHD, coloc subsets for systolic BP and BMI versus T2D. In certain cases, pronounced differences were only observed in a single setting using one model type but not the others, such as: coloc-based partitioning for BMI with respect to CHD (1 sample multivariable), Mendelian disease-based partitioning of DBP and SBP with respect to heart failure (2 sample univariable). In the first instance, one may expect for some differential effect to become apparent only in the multivariable analysis when adjusting for the shared effect. Nevertheless, lack of effect difference replication in corresponding two-sample MVMR is troubling, as the instrumental variable subsets were found not to suffer from weak instrument bias previously^12^. The second example involving effect difference observed only in two-sample univariable analysis should be treated as less reliable due to poor replication in one-sample setting, especially in multivariable analysis evaluating direct effects.

Therefore, to gain a better understanding of tissue- or pathway-partitioned effects multiple types of MR analyses should be undertaken, whenever possible. For instance, in a recent publication, Leyden et al. (2023)^12^ applied the coloc method to study the effect of adipose and brain tissues in BMI on cancer in two-sample setting only, due to availability of one-sample datasets with sufficiently large numbers of cases. While they noted that adipose-derived variants may predominantly drive the association with endometrial cancer, our random sampling control found weak support for a significant difference between adipose and brain in a multivariable analysis (p-value=0.161).

We expected that stratification of SNPs by pathway and tissues should result in reduced heterogeneity compared to using all SNPs, which can capture a wider range of biological processes. However, our coloc- and Mendelian disease-based results did not consistently support this expectation. For some SNP subsets, the heterogeneity was indeed lower, which was in line with the initial hypothesis.

However, other SNP subsets showed greater heterogeneity than all SNPs combined, suggesting that those specific pathways or tissues might still be highly pleiotropic or indicate presence of some bias in the method, such as misclassification of SNP functional category.

Our new approach uses enrichment of ontology terms assigned to Mendelian diseases whose causal genes share the same genetic locus as the exposure SNPs, which can sometimes result in ambiguous assignment to appropriate biological pathways. For example, rs12630999 from BMI was allocated to both the “mental health” and “metabolic” set due to location between two neighbouring Mendelian disease genes: *STAG1* and *PCCB*, respectively. In another case, one systolic BP variant (rs3915499) was associated with two different pathways (“renal” and “vessel”) due to the two distinct monogenic diseases caused by disruption of smooth muscle myosin heavy-chain 11 gene^71^ whose intron the SNP resides in. Under such a scenario, MendelVar cannot provide a more nuanced prediction regarding the “correct” pathway(s). In this example, “vessel”-only assignment could be more suitable as the associated disease, familial thoracic aortic aneurysm 4 is defined by profound structural abnormalities in the aorta, while a renal symptom (hydronephrosis) is only a marginal feature of megacystis-microcolon-intestinal hypoperistalsis syndrome 2. Nonetheless, we found that in many cases MendelVar unequivocally assigns exposure SNPs to genes causal for disorders with strong links to a single category: “renal” - *CEP164* (nephronophthisis 15), *NRIP1* and *PBX1* (congenital anomalies of kidney and urinary tract syndromes); “vessel” - *EIF2AK4* (familial pulmonary capillary hemangiomatosis), *PRDM6* (patent ductus arteriosus), *HTRA1* (cerebral arteriopathy with subcortical infarcts and leukoencephalopathy); “mental health” - *PPP3CA* (developmental and epileptic encephalopathy 91, ACCIID), *KCNMA1* (cerebellar atrophy, developmental delay, and seizures, Liang-Wang syndrome); “metabolic” – *PPARG* (familial lipodystrophy), *SLC2A2* (Fanconi-Bickel syndrome). Correct SNP assignment to the gene in each case was also supported by either presence within its intron or coding region.

Similarly to Darrous et al. (2023)^9^, we conclude that our new method offers complementary, orthogonal SNP stratification to the existing colocalisation approach as evidenced by very weak overlap. Comparison of MR results using both approaches suggests agreement for the dominant role of vasculature-related SNPs in determining the left ventricular stroke volume and the risk of type 2 diabetes, but discordance with regards to coronary heart disease, and influence of different BMI pathways on atrial fibrillation. It is not straightforward to identify the cause of discrepancy. While coloc-based gene selection shows poor enrichment of relevant gene functional categories in general (STE 21-24) unlike Mendelian-based selection, the latter gene categorisation is based on disease symptom ontologies which are a-priori more directly related to gene function than expression pattern. We find that more Mendelian “metabolic” than “mental health” SNPs are shared with coloc “brain” SNPs which could be partially due to weak evidence for involvement of coloc “brain” genes in neurodevelopmental pathways and/or brain-centric expression of some of the key genes regulating metabolism^72,73^. Furthermore, although both Mendelian “metabolic” and coloc “adipose” subsets are enriched for metabolic pathway genes, the “metabolism” term is broad and there could still be stark differences in proportion of metabolic pathways represented in each subset. Disease Ontology used for assignment of genes to Mendelian disease categories for BMI is also quite sparsely annotated as indicated by reduction in the number of pathway-assigned SNPs relative to blood pressure where Human Phenotype Ontology was used; that may well have introduced selection bias.

While this study introduced and evaluated a novel, Mendelian disease-centric approach to dissecting the impact of different biological pathways on complex risk factors in MR, several issues warrant further investigation. Applying the method to other complex traits and physiological pathways could result in more nuanced understanding of shared genetic risk factors in cross-disease analyses. Mendelian disease-partitioning method is reliant on mainly manual curation of disease ontology terms based on descriptions of clinical features^64,66^, and expanding the automation of that time-consuming process could increase power and potentially accuracy of our method. The specificity of the colocalisation method suffers from co-ordinated expression of genes across many tissues which complicates selection of biologically causal tissues over merely tagging but new methods are being developed to address this confounding factor^74,75^. Furthermore, we emphasise that the modest eQTL sample sizes analysed in this study may have reduced the number of instruments we were able to identify with robust colocalization evidence^76^. Next, integrating different features used for exposure partitioning into a single joint model may offer improvement in pathway-based stratification. Here, we used Mendelian and gene expression information but other orthogonal evidence such as protein-protein interactions^77^ and PheWAS^9^ could be combined. Such (and other) future new methods would be useful to address discordance in the dominant biological pathways identified for certain disease outcomes between coloc- and Mendelian disease- based instrument partitioning.

## Author contributions

MKS: Conceptualization, Methodology, Formal analysis, Visualization, Writing – original draft, Writing – review and editing

TGR: Resources, Writing - review and editing GML: Resources, Writing - review and editing

TG: Supervision, Methodology, Resources, Writing – review and editing, Funding acquisition

## Data and code availability

GWAS summary statistics availability is provided by relevant publications. We accessed the following summary statistics via OpenGWAS: ebi-a-GCST006414, ebi-a-GCST006906, ebi-a-GCST009541, ieu-a-7, ieu-a-26, ieu-a-798, ieu-b-38, ieu-b- 39, ieu-b-40.

## Supporting information

Supplementary Dataset STE

## Data Availability

GWAS summary statistics availability is provided by relevant publications. We accessed the following summary statistics via OpenGWAS: ebi-a-GCST006414, ebi-a-GCST006906, ebi-a-GCST009541, ieu-a-7, ieu-a-26, ieu-a-798, ieu-b-38, ieu-b-39, ieu-b-40.

## Acknowledgements

This research has been conducted using the UK Biobank Resource under Application Number 81499.

## Ethics approval

UK Biobank received ethical approval from the Research Ethics Committee (REC reference: 11/NW/0382).

