## Supplementary Figures for "Distinct pathway-based effects of blood pressure and body mass index on cardiovascular traits: comparison of novel Mendelian Randomization approaches"

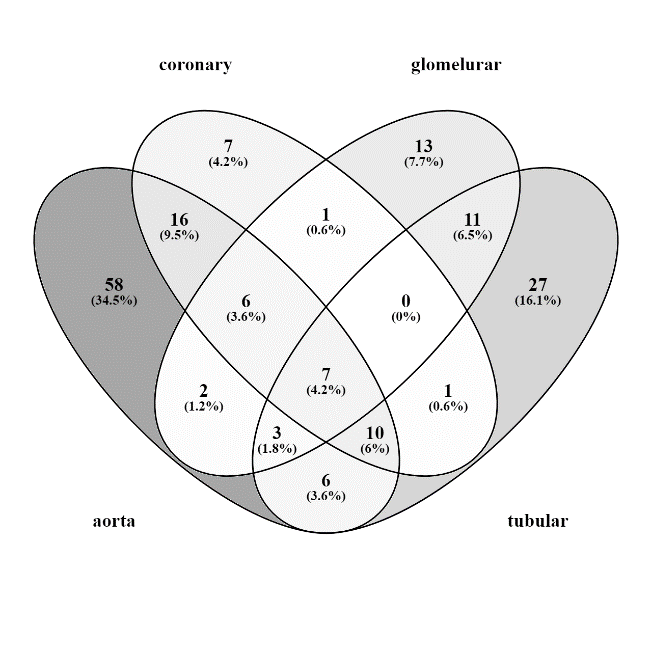

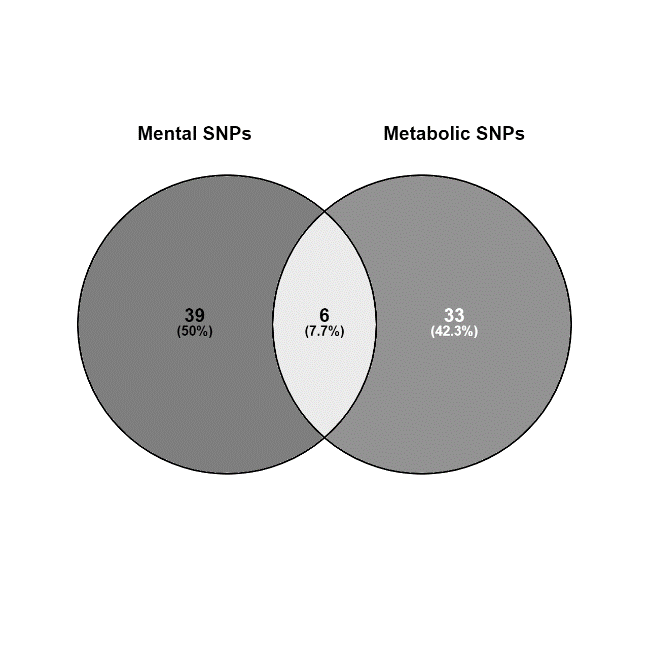

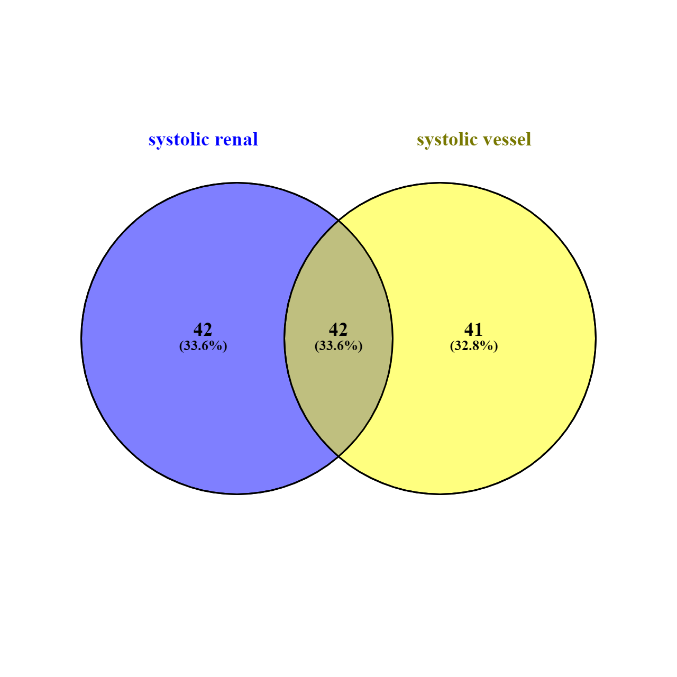

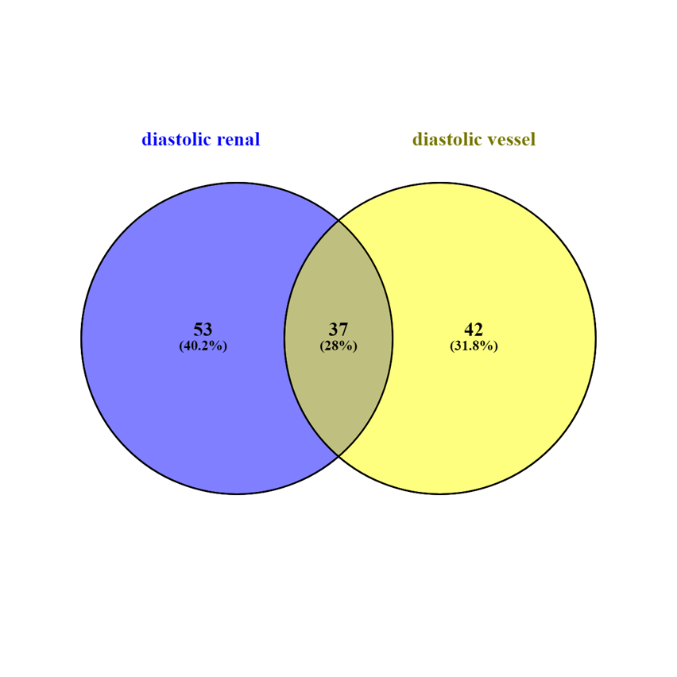

**C**

**D**

**B**

**A**

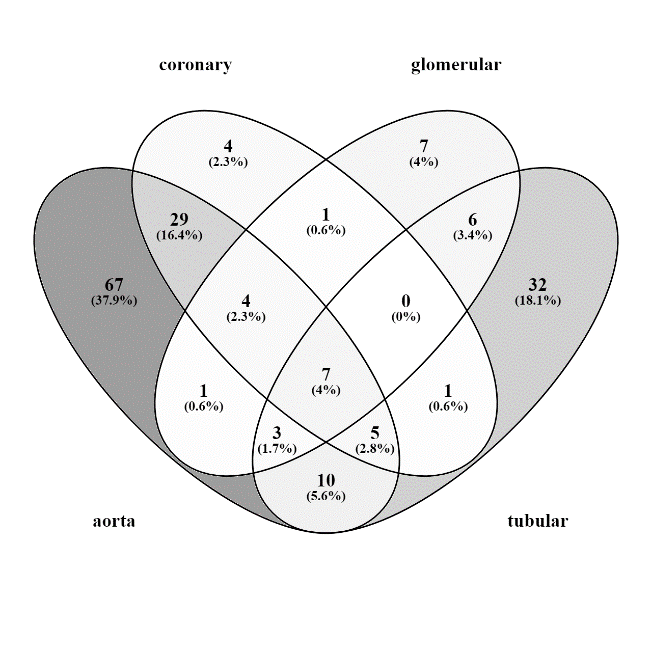

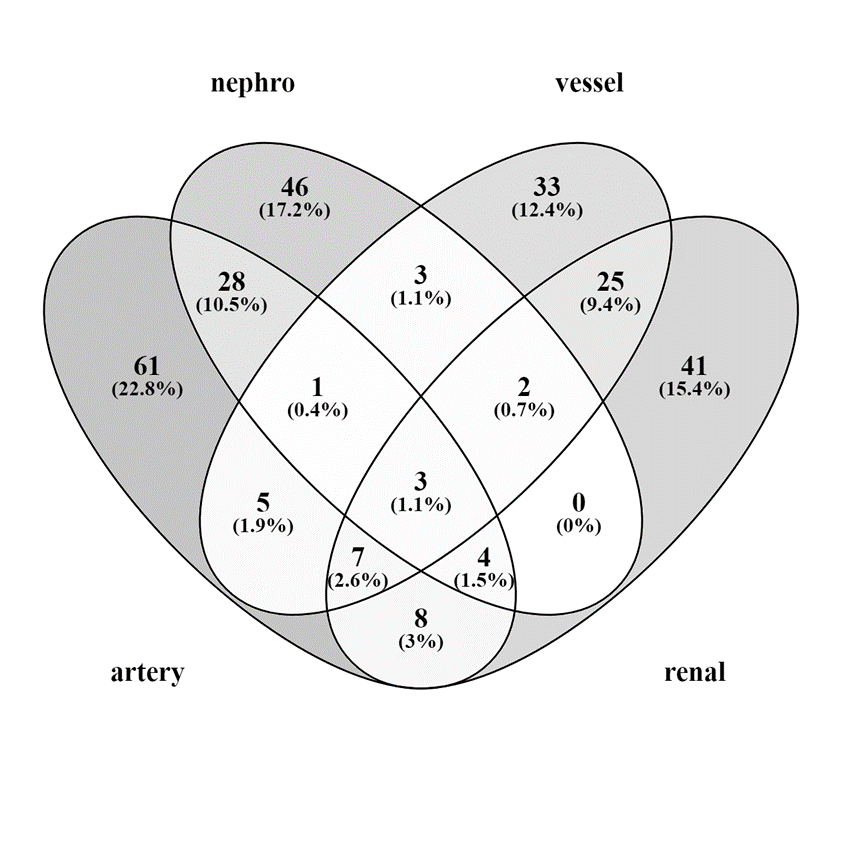

**E**

**F**

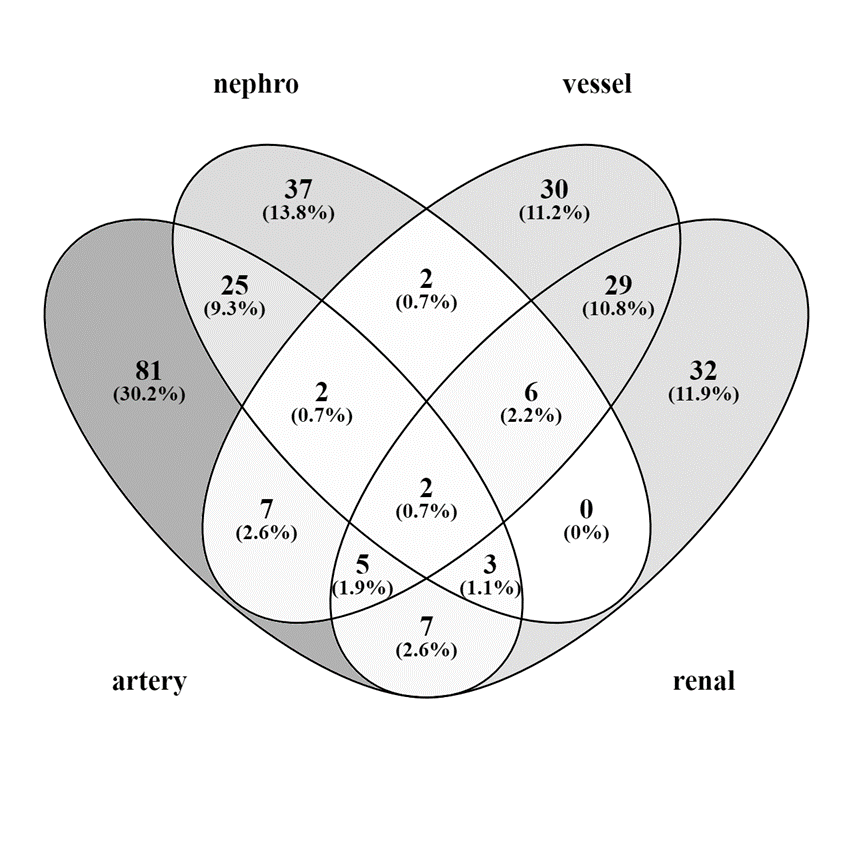

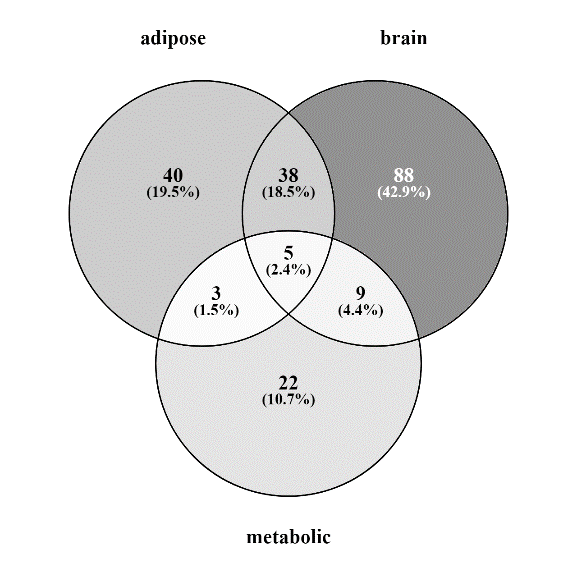

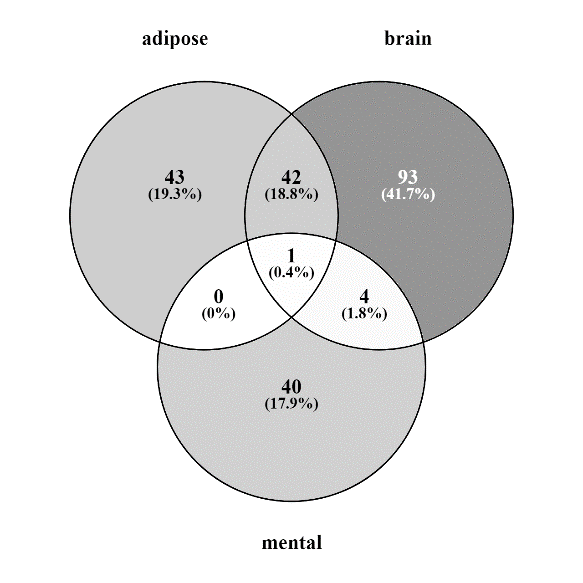

**H**

**I**

**G**

**Supplementary Figure 1. SNP overlap of Mendelian and coloc subsets.**

A) Mendelian diastolic BP SNPs – renal and vessel; B) Mendelian systolic BP SNPs – renal and vessel; C) Mendelian BMI SNPs – mental and metabolic; D) coloc diastolic BP SNPs – artery (aorta, coronary) and nephro (glomerular and tubular); E) coloc systolic BP SNPs – artery (aorta, coronary) and nephro (glomerular and tubular); F) Mendelian diastolic BP SNPs – renal and vessel vs coloc diastolic BP SNPs – artery and nephro; G) Mendelian systolic BP SNPs – renal and vessel vs coloc diastolic BP SNPs – artery and nephro; H) Mendelian BMI SNPs – mental vs coloc BMI SNPs – adipose and brain; I) Mendelian BMI SNPs –metabolic vs coloc BMI SNPs – adipose and brain. In H) and I), comparison between coloc and Mendelian SNP subsets was made not using just original SNPs but also their proxies (r^2^ > 0.8, 1 Mbp, 1000 Genomes European population) since the coloc SNP dataset contained additional SNPs from UKBB BMI GWAS.

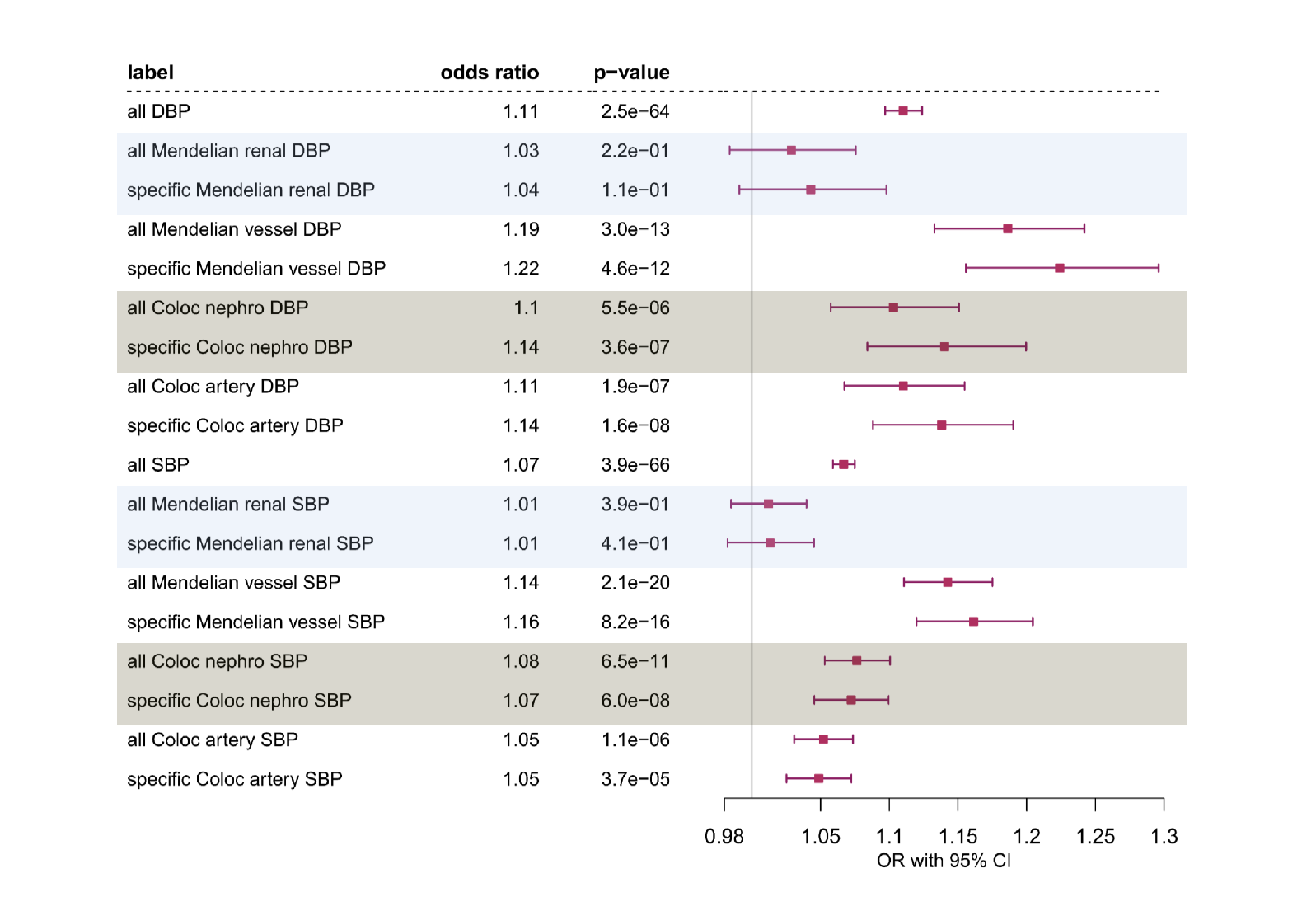

**Supplementary Figure 2. Myocardial infarction:** one sample Multivariable Mendelian Randomization analysis of the effect of diastolic blood pressure (DBP) and systolic blood pressure (SBP) on MI using all SNPs, all/specific Mendelian disease partitioned (disease with abnormalities in the renal or blood vessel system) genetic instruments and all/specific Coloc partitioned (nephro – kidney tissues: glomerular and tubulointerstitial, artery – aorta and coronary artery tissues) instruments. Effect sizes are scaled to per one SD change in blood pressure.

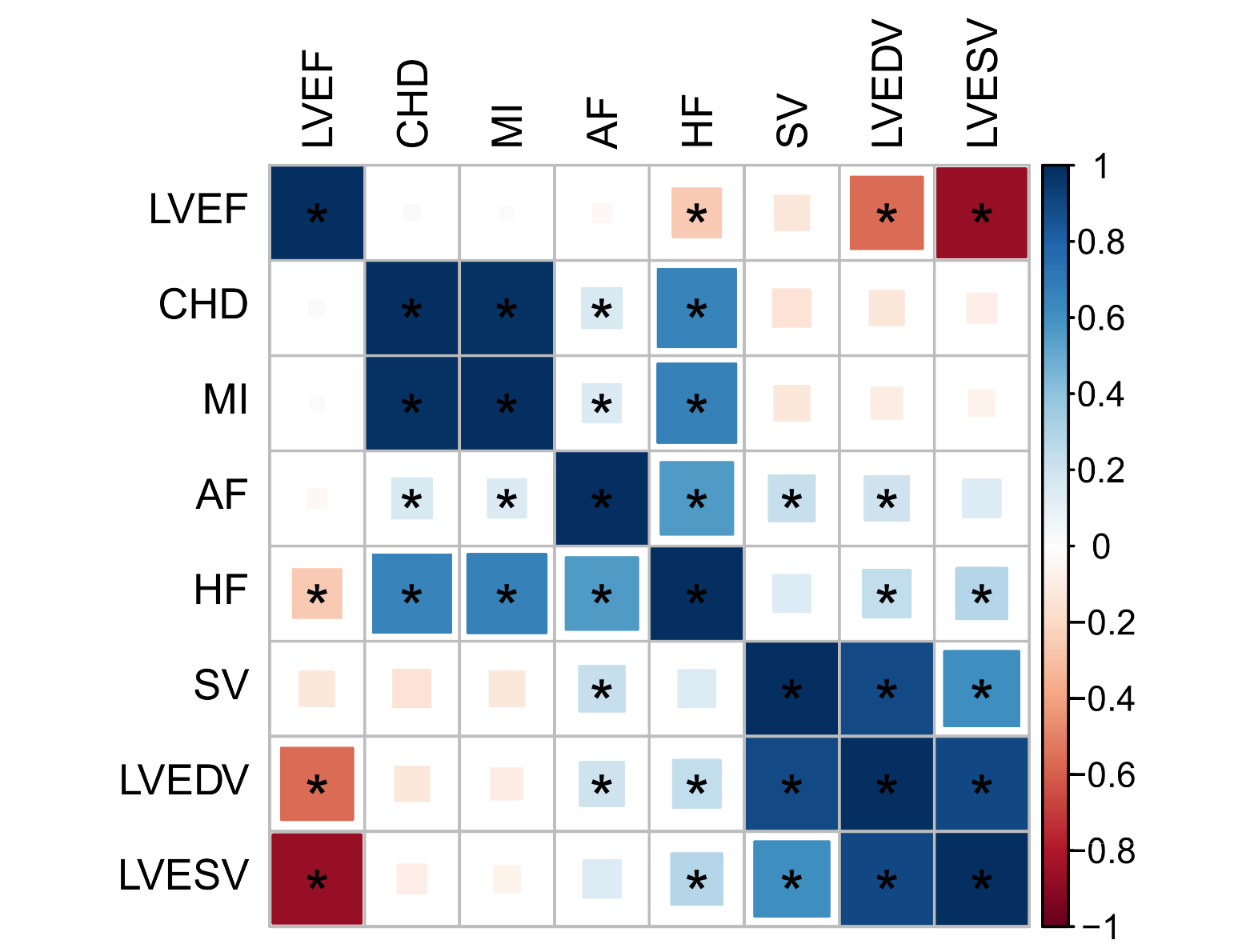

**Supplementary Figure 3.** Pairwise genetic correlation among cardiac traits estimated using LD score regression in LDSC. Larger area of square filling corresponds to more significant FDR and significant correlation coefficients (FDR < 0.05) are indicated by asterisks. AF - atrial fibrillation, HF - heart failure, CHD - coronary heart disease, MI - myocardial infarction, SV - left ventricle stroke volume, LVEF - left ventricular ejection fraction, LVEDV - left ventricular end-diastolic volume, LVESV - left ventricular end-systolic volume.

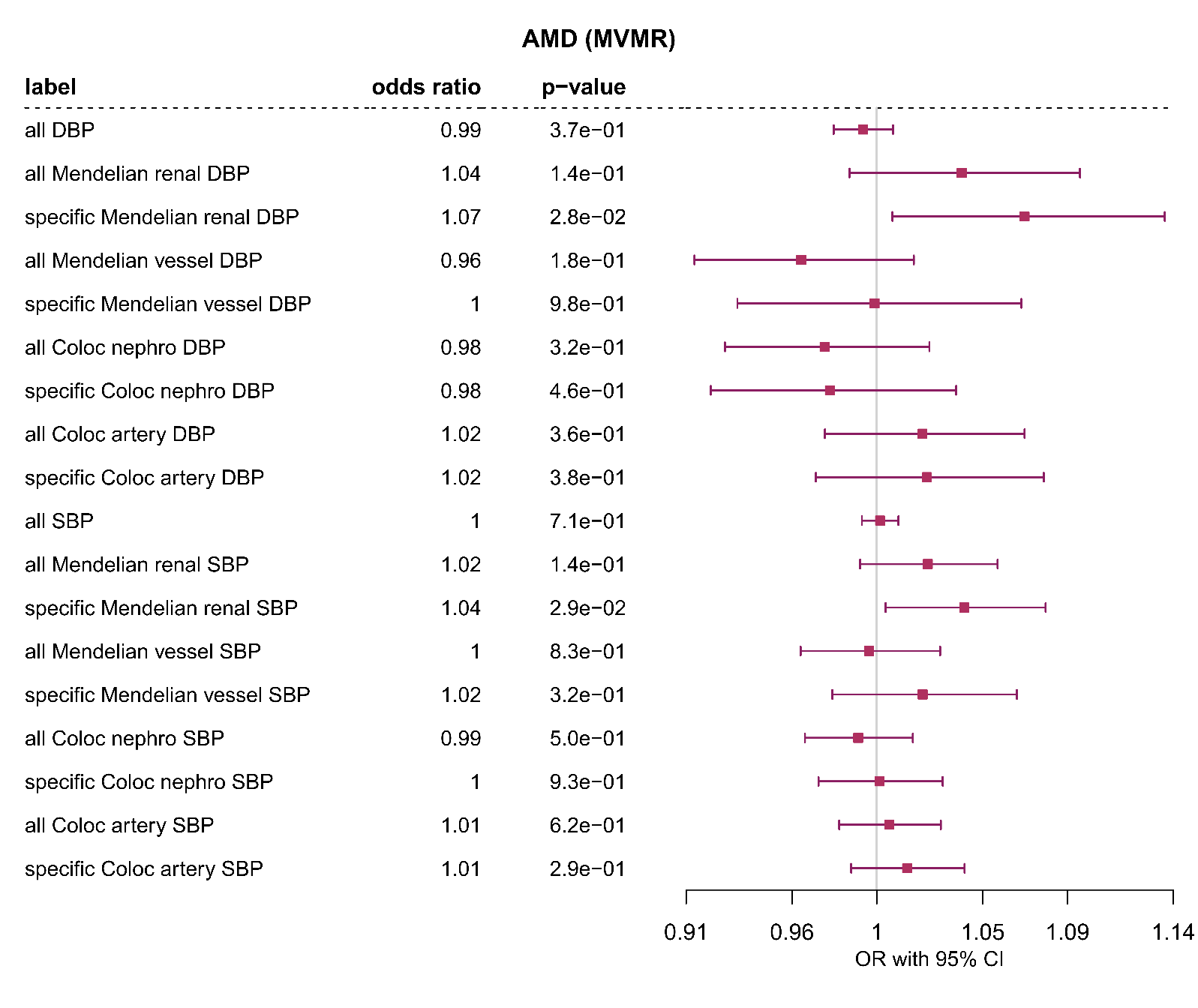

**Supplementary Figure 4.** **Age-related macular degeneration:** One sample Multivariable Mendelian Randomization analysis of the effect of diastolic blood pressure (DBP) and systolic blood pressure (SBP) on age-related macular degeneration (AMD) using all SNPs, all/specific Mendelian disease partitioned (disease with abnormalities in the *renal* or blood *vessel* system) genetic instruments and all/specific Coloc partitioned (*nephro* – kidney tissues: glomerular and tubulointerstitial, *artery* – aorta and coronary artery tissues) instruments. Effect sizes are scaled to per one SD change in blood pressure.

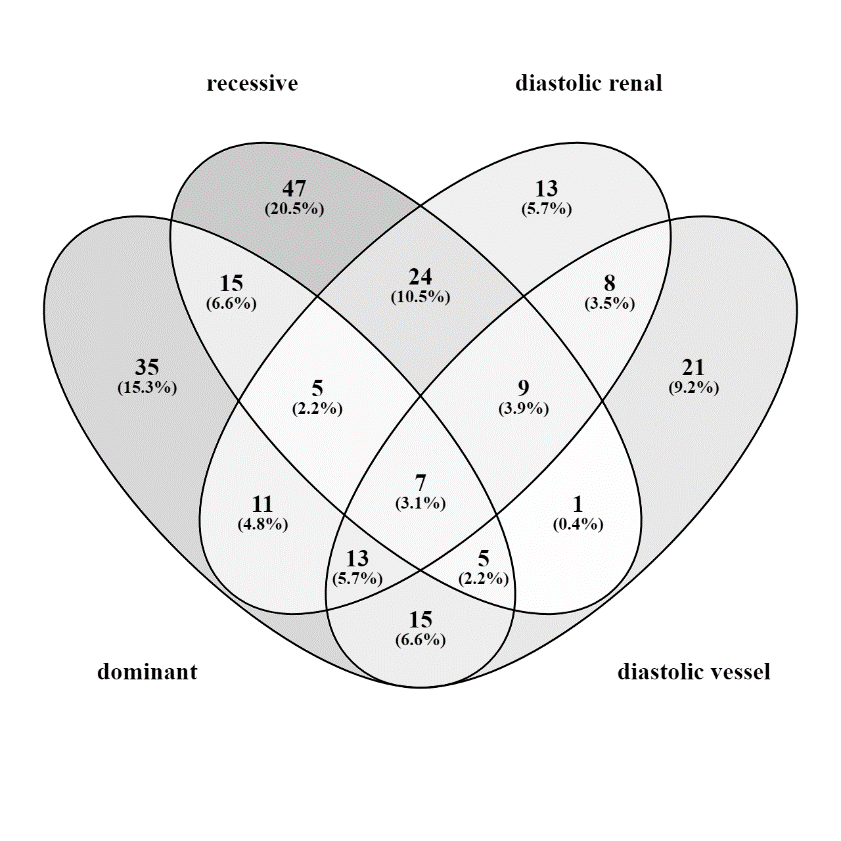

**A**

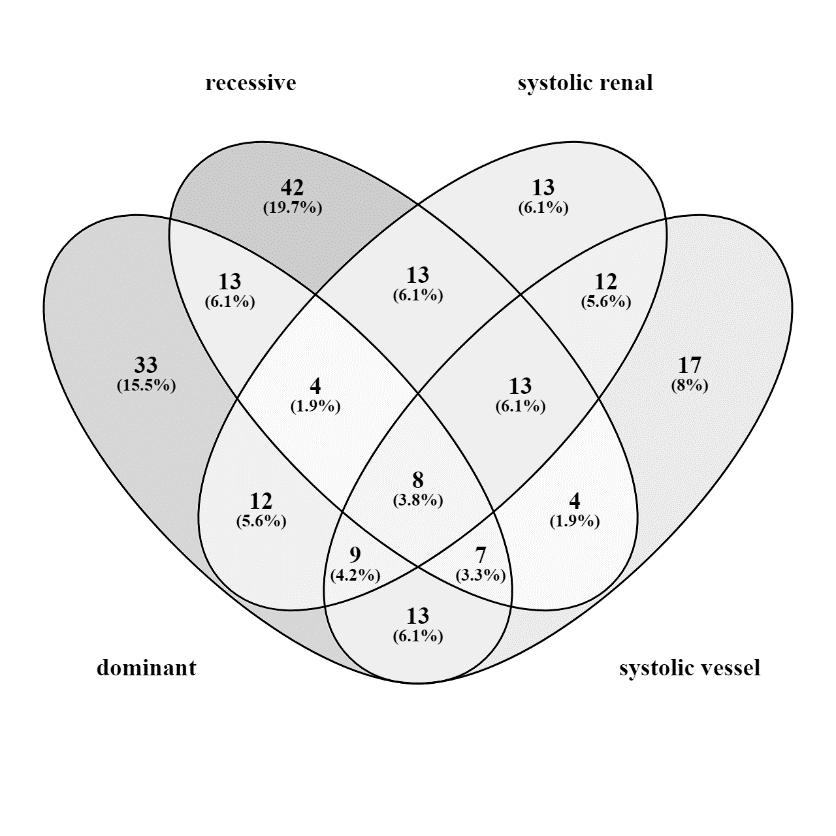

**B**

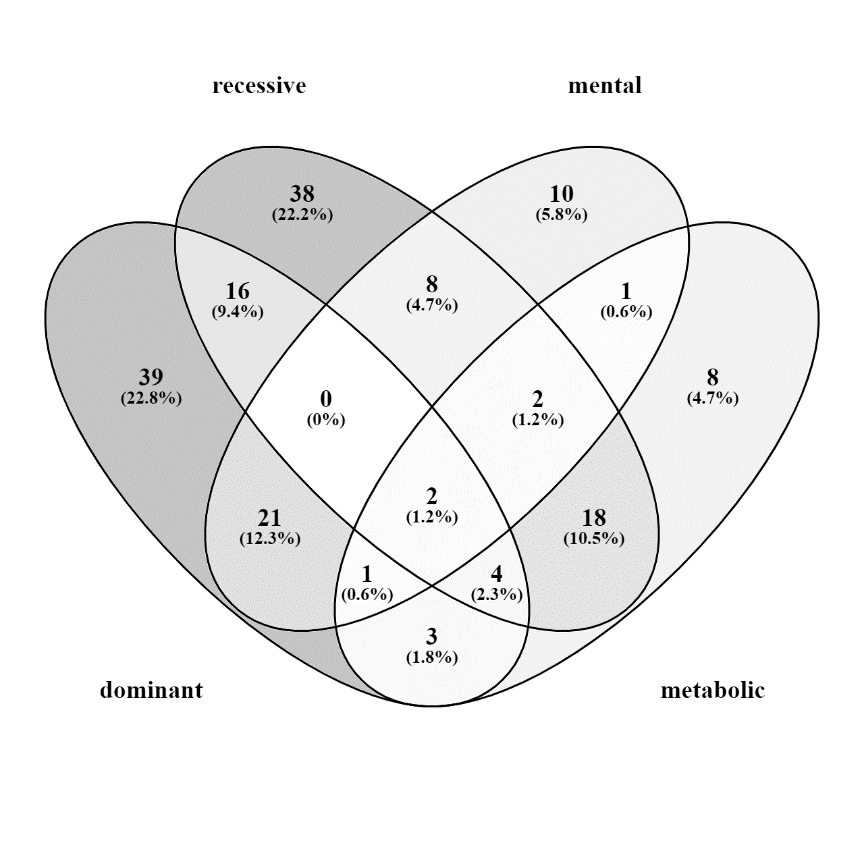

**C**

**Supplementary Figure 5.** Comparison of SNPs associated with enriched Mendelian disease terms and SNPs associated with Mendelian disease genes inherited in autosomal dominant and autosomal recessive fashion: A) diastolic blood pressure; B) systolic blood pressure; C) body mass index.

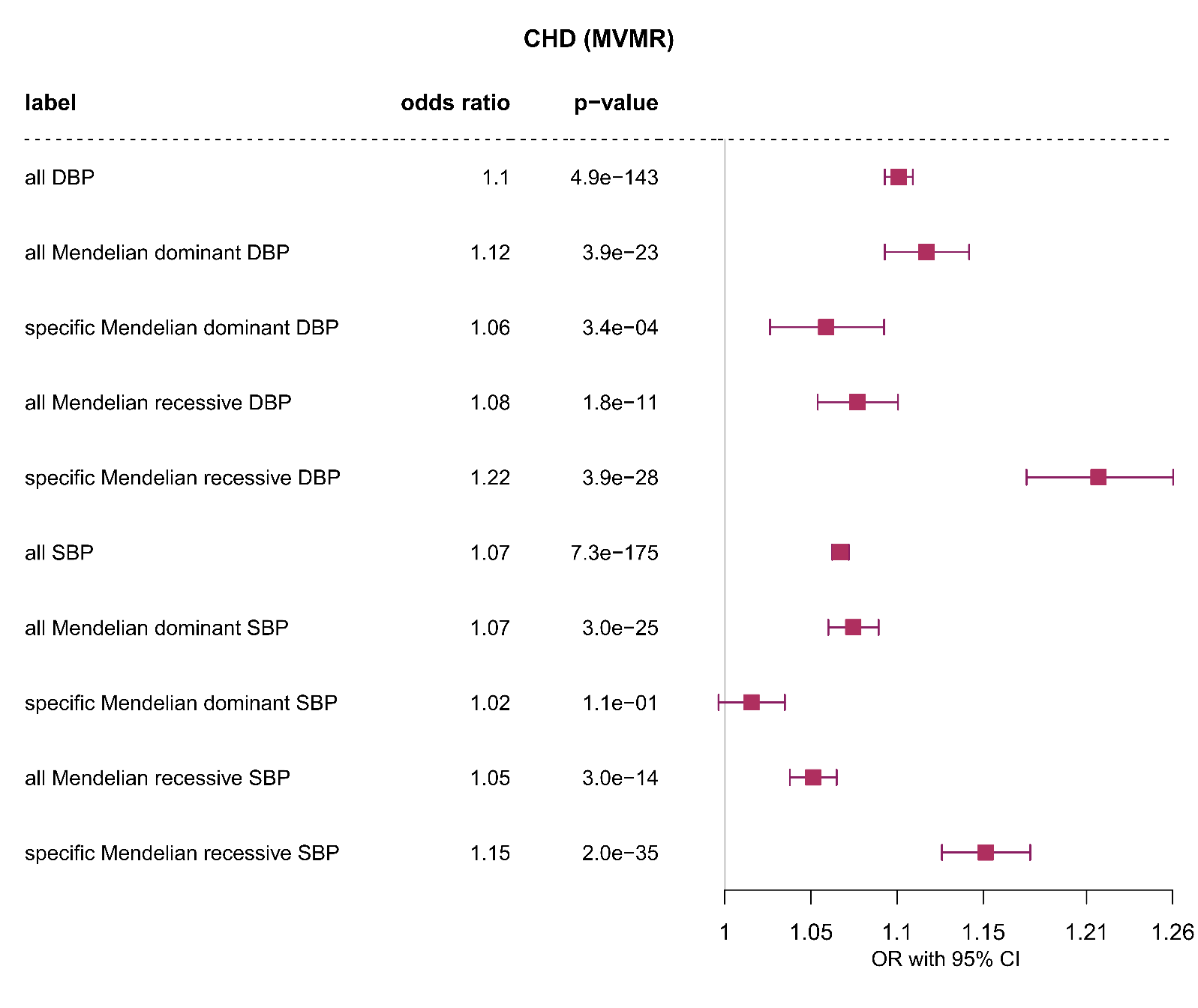

**Supplementary Figure 6. Coronary heart disease:** One sample Multivariable Mendelian Randomization analysis of the effect of diastolic blood pressure (DBP) and systolic blood pressure (SBP) on CHD using all SNPs, all/specific Mendelian disease-partitioned (disease with autosomal *dominant* or *recessive* inheritance pattern) genetic instruments. Effect sizes are scaled to per one SD change in blood pressure.

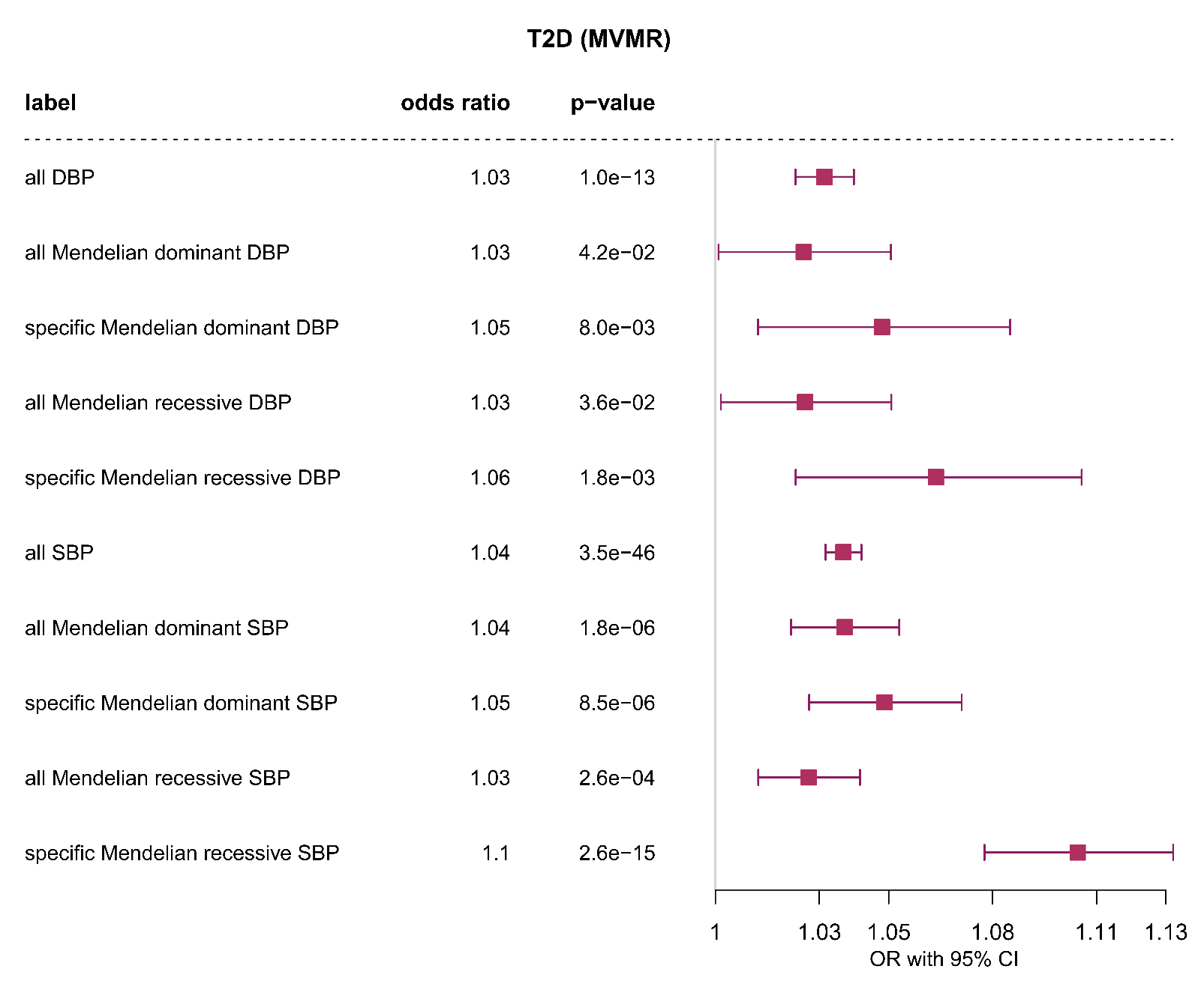

**Supplementary Figure 7. Type 2 diabetes:** One sample Multivariable Mendelian Randomization analysis of the effect of diastolic blood pressure (DBP) and systolic blood pressure (SBP) on T2D using all SNPs, all/specific Mendelian disease-partitioned (disease with autosomal *dominant* or *recessive* inheritance pattern) genetic instruments. Effect sizes are scaled to per one SD change in blood pressure.
